# Compliance with Covid-19 measures: evidence from New Zealand

**DOI:** 10.1101/2021.04.08.21255157

**Authors:** Geoff Kaine, Suzie Greenhalgh, Vic Wright

## Abstract

Governments around the world are seeking to slow the spread of Covid-19 by implementing measures that encourage, or mandate, changes in people’s behaviour. These changes include the wearing of face masks, social distancing, and testing and self-isolating when unwell. The success of these measures depends on the commitment of individuals to change their behaviour accordingly. Understanding and predicting the motivation of individuals to change their behaviour is therefore critical in assessing the likely effectiveness of these measures in slowing the spread of the virus.

In this paper we draw on a novel framework, the I_3_ Compliance Response Framework, to understand and predict the motivation of residents in Auckland, New Zealand, to comply with measures to prevent the spread of Covid-19. The Framework is based on two concepts. The first uses the involvement construct to predict the motivation of individuals to comply. The second separates the influence of the policy measure from the influence of the policy outcome on the motivation of individuals to comply.

In short, the Framework differentiates between the strength of individuals’ motivation and their beliefs about the advantages and disadvantages of policy outcomes and policy measures. We found this differentiation was useful in predicting an individual’s possible behavioural responses to a measure and discuss how it could assist government agencies to develop strategies to enhance compliance.

## Introduction

The success of measures to slow or stop the spread of Covid-19 such as wearing face masks and social distancing, depends on the commitment and capacity of individuals to comply with them, and change their behaviour accordingly [1–3]. Ineffective compliance with these measures can put the achievement of policy outcomes at risk [4,5]. For example, failure to wear face masks and socially distance may put the outcome of eliminating Covid-19 from countries such as New Zealand at risk and may mean considerable resources must be invested in enforcement to avoid increased rates of infection, higher mortality, and the imposition of lockdowns causing both economic and psychological damage. Hence, understanding and predicting the extent to which individuals are motivated to change their behaviour to comply with measures is critical in assessing how effective these measures are likely to be, and whether alternatives such as curfews and lockdown can be avoided.

In this paper we draw on a novel framework, the I_3_ Response Framework [6], to understand and predict the motivation of residents of Auckland, the largest city in New Zealand, to comply with measures to prevent the spread of Covid-19, such as the wearing of face masks, self-isolating when unwell, and getting tested for Covid-19. Auckland is the port of entry for many people coming to New Zealand and therefore at high risk of frontline border workers being exposed to the virus. The Framework is applied to model the engagement with, and beliefs and feelings about, government measures proposed or taken to deal with some policy outcome. The target group is composed of those whose acceptance of the measures, which is not inevitable, is essential for the achievement of the desired policy outcome. Our aim was to test the proposition that compliance with policy measures to prevent the spread of Covid-19 depends on people’s involvement with the measures (how much an individual cares about them) as well as their attitude towards them.

Fundamentally, the Framework differentiates between the strength of individuals’ motivation and their beliefs about the advantages and disadvantages of policy outcomes and policy measures. We found this differentiation was useful in predicting an individual’s possible behavioural responses to a measure, which could assist government agencies to develop strategies to enhance compliance. With respect to preventing the spread of Covid-19 in New Zealand we found that Auckland residents’ willingness to wear face masks and self-isolate if unwell depended on the strength of their involvement (motivation) with, as well as their attitudes towards, both the policy outcome (preventing the spread of Covid-19) and the policy measures themselves (wearing face masks and self-isolating if unwell).

The results highlight the importance of distinguishing unintentional non-compliance with respect to wearing face masks, self-isolating when unwell, and testing from deliberate non-compliance, and tailoring enforcement strategies appropriately. The results also highlight the difficulty of communicating effectively through mass media with those who have low involvement with preventing the spread of Covid-19. They draw attention to the importance of distinguishing between those with low and high involvement in considering the possible effects on compliance of the dissemination of misinformation about Covid-19 through social media.

## Theory

Kaine et al. [6] proposed that theories about people’s responses to policy measures have a common underpinning, whether grounded in the economics of rational choice or more behavioural, such that people’s decision-making is motivated by the achievement of personal goals, and the importance of a decision to an individual influences the extent to which they will devote cognitive effort to gathering information, processing that information, formulating attitudes, and reaching a decision. Kaine et al. [6] suggested that these theories cannot be expected to predict behaviour when a decision is not perceived to be important enough (i.e. sufficiently relevant to people’s personal goals) to trigger the effort required to form an attitude that has the power to influence their behaviour. Consequently, to predict how people may or may not respond to any given policy measure, it is necessary to understand whether they are likely to invest effort in decision-making regarding that measure.

As explained in detail in Kaine et al. [6], the effort people will devote to decision-making about complying with a policy measure will depend on their *involvement* with the policy *issue* (in this case the policy outcome of eliminating Covid-19 from New Zealand) and the *intervention* (the policy measure, such as wearing face masks, self-isolating if unwell and being tested for Covid-19), with the former being an important component of the context for the latter. These concepts underpin the I_3_ Framework^1^ used in this analysis.

In a specific applied setting, such as a policy to eliminate Covid-19, the I_3_ Framework enables the prediction of likely compliance with policy measures, and given the reasons for their involvement and their attitudes, the best ways to enhance that compliance. The behaviour changes to be analysed with the I_3_ Framework occur in a public policy context rather than a commercial context. This means the outcome(s) sought are typically declared, and government, or its agencies, intervene with measures designed to modify behaviour in pursuit of the outcome(s). Either compulsion or voluntary responses may be involved, but compliance is central to achieving outcomes. In what follows in this section we have drawn extensively on the discussion of the interpretation of Framework findings from Kaine et al. [6] to make it readily accessible for the reader.

### The I_3_ Framework

Kaine et al. [6] proposed that people’s responses to policy measures, such as the requirement to wear face masks, can be inferred from their:

- involvement with the relevant policy outcome (preventing the spread of Covid-19)
- involvement with and attitude towards the policy measure itself (e.g. wearing face masks).

Involvement with the policy measure signals the degree to which the measure itself is a source of motivation for the individual, irrespective of the policy issue [7,8]. This allows for the possibility that individuals are motivated to act in response to a measure even though they do not perceive the policy outcome the measure addresses is relevant to them. In such situations, it may be that the wish to comply is motivated by involvement with some other outcome, such as achieving perceived conformity with social norms [9].

However, the perceived relevance of a policy outcome is plainly relevant to an individual’s cognitions about related measures. One would expect a positive correlation between involvements with each. The value of the Framework is that it enables a decomposition of overall involvement with a policy outcome and corresponding measures, as well as distinguishing between involvement with different measures and closer analysis of the role of beliefs held by individuals, as informational contexts for attitudes.

The two dimensions of involvement with the policy outcome and involvement with the policy measure mean that the responses of people to a policy measure can be classified into four quadrants, as shown in Fig 1.

**Fig 1.**
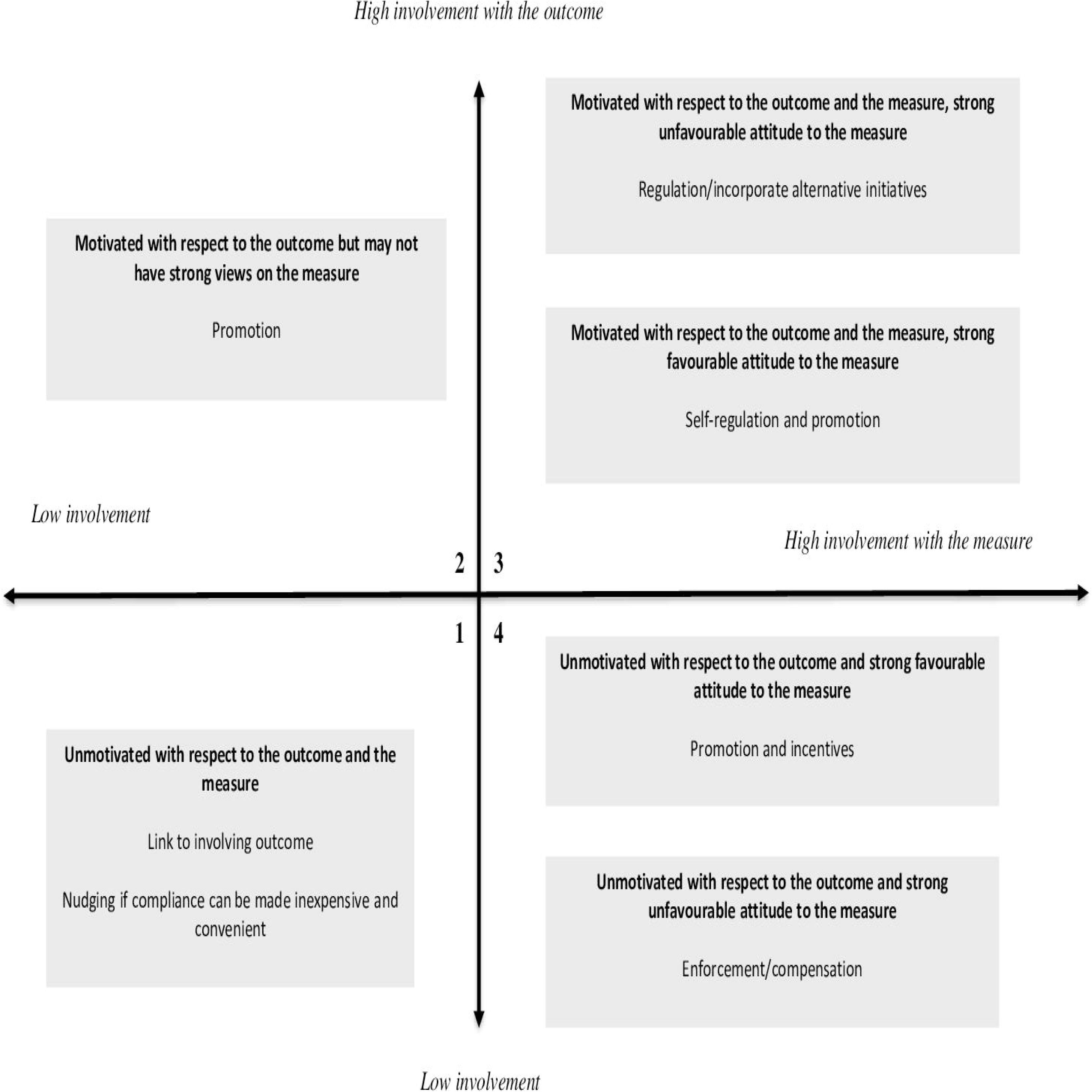
Map and explanation of quadrants in the I_3_ framework.

People in quadrant 1 exhibit low involvement in both the policy outcome and the policy measure. These people are likely to have little knowledge, or even awareness, of the policy outcome and are likely to have limited knowledge of the policy measure, and to have weak attitudes towards it, if any at all. In terms of Kassarjian [10], these people may appear to be either detached (i.e. they have other interests and concerns) or ‘know-nothings’ (people who do not particularly care about or have no interest in the outcome). Non-compliance with the measure is largely unintentional [11]. Chaffee and Roser [12, p. 376] describe their behaviour as being ‘a direct response to situational constraints and not especially reflective of one’s attitudes or knowledge.’

If people in quadrant 1 present little risk in terms of achieving aggregate policy outcome targets, they can be ignored [6]. Otherwise, their compliance with the measure may be encouraged by:

- linking the policy outcome to a subject they find more involving
- reducing the effort required to be compliant
- promoting awareness of the policy outcome and the policy measure.

However, because people in this quadrant are disinterested, they are unlikely to pay attention to promotional messaging, hence the last strategy of promoting awareness is likely to be ineffective. Kim [7] suggests that an affect-evoking strategy (i.e. one that evokes an emotional response) should be the most effective means of attracting attention under these circumstances. This is most likely to be achieved by focusing on the policy outcome as low involvement, with the outcome acting as a hurdle to greater involvement with individual measures.

People in quadrant 2 exhibit high involvement with the policy outcome but low involvement with the measure. Consequently, they would be aware of the outcome and invest time and energy in processing information, decision-making and responding to the outcome [12,13]. They may have limited knowledge of the policy measure and may have weak or ambiguous attitudes towards it. Any non-compliance with the measure is largely unintentional [6].

If people in quadrant 2 represent little risk in terms of achieving the policy outcome they can be ignored. If their compliance is important to achieving the policy outcome, reducing the effort required for compliance [14] and promoting awareness of the policy measure may be worthwhile by taking advantage of the intensity of their involvement with the policy outcome, particularly when this is accompanied by favourable attitudes.

People in quadrant 3 exhibit high involvement with both the policy outcome and the measure. These people are likely to have extensive and detailed knowledge of the policy outcome. They are also likely to have extensive knowledge of the policy measure and strong attitudes towards it [13]. If their attitude towards the policy measure is favourable, they will comply with the measure and may even advocate for it [11]. Consequently, a strategy for promoting compliance among individuals in this quadrant with a favourable attitude might focus on self-regulation, using mechanisms such as voluntary codes of conduct. Promotion and monitoring may also be worthwhile to ensure awareness and knowledge of obligations, ensure desirable behaviours are maintained, and identify at an early stage any changes in their attitude [6].

If people in quadrant 3 have an unfavourable attitude towards the policy measure, they may comply, but reluctantly [6]. Non-compliance with the measure will be intentional. Most likely they will prefer – and even advocate for – changes to the design of the policy measure. Where practical, incorporating these changes may encourage the compliance of these people [15]. Alternatively, offering incentives to reduce compliance costs may neutralise unfavourable reactions.

Another strategy for promoting compliance among individuals in this quadrant with an unfavourable attitude is to change their attitude towards the measure. This may be possible by reframing the benefits of the measure in terms of another, more involving subject [6], thus provoking a recalculation of net costs and benefits. Alternatively, a promotional programme may be implemented with the outcome of persuading these individuals they are mistaken, and that the behaviours required by the policy measure are superior to any alternatives. Finally, compliance among these individuals might be increased by investing resources in enforcement, to increase the likelihood of detection and prosecution, and legislating severe penalties for non-compliance.

Note that if the causes of non-compliance relate to unpredictable variations in the environment, or to unforeseeable technical problems, then enforcement and general deterrence may be ineffective. A more appropriate strategy in these circumstances may be to focus on the provision of technical assistance [16,17].

People in quadrant 4 exhibit low involvement with the policy outcome but high involvement with the measure. People in this quadrant are likely to have limited knowledge of the policy outcome. They are likely to have detailed knowledge of the policy measure and have strong attitudes towards it [13]. If their attitude towards the measure is favourable, then they will comply with the measure [6]. In these circumstances, the government agency may play a monitoring role to check the conditions promoting compliance do not change. A promotional strategy to support and reinforce compliance behaviour may also be worthwhile.

On the other hand, if the members of this quadrant have an unfavourable attitude towards the policy measure, they will only comply reluctantly, or may intentionally refuse to comply at all. These people will regard the measure as imposing unwarranted costs upon them. Most likely they will agitate against the policy measure [6] because they are not committed to the outcome. One strategy for promoting compliance among these individuals is to change their attitude towards the measure. This may be possible by reframing it in terms of another, more involving subject [6]. Offering incentives to offset compliance costs, or delaying or staging the introduction of policy measures, may neutralise unfavourable reactions [15]. Finally, compliance among these individuals might be increased by investing resources in enforcement to increase the likelihood of detection and prosecution of, and by introducing severe penalties for, non-compliance.

In summary, Kaine et al. [6] hypothesised that individual responses to policy measures will depend on the intensity and source of involvement of the individual with the measure and, where that involvement is sufficiently intense to form an attitude, on whether that attitude is favourable or unfavourable. The I_3_ Framework has been employed to understand and predict compliance behaviour in a variety of contexts in agriculture [16,18,19,20], rural and urban predator control [21,22], and community support for predator control [23]. The results of these studies suggests the Framework has predictive and differential validity with respect to compliance behaviour and discriminant validity with respect to attitudes.

### Covid-19 in New Zealand

Covid-19 was first detected in New Zealand on 28 February 2020 [24]. Within three weeks the central government had closed New Zealand’s international border to all except returning citizens and permanent residents. The government began pursuing a restrictive strategy [25] of eliminating Covid-19 and applied a range of control measures to stop the transmission of Covid-19 in New Zealand [26]. Elimination did not necessarily mean eradicating the virus permanently from New Zealand; rather, that central government was confident chains of transmission in the community had been eliminated for at least 28 days, and any cases imported from overseas in the future could be effectively contained [26].

The central government instituted a four-tier alert system that mandated measures such as: progressively tighter restrictions on people’s movement outside their homes and immediate families, including travelling to work; social distancing and encouraging the wearing of masks outside the home at the higher alert levels; and self-isolating and seeking testing if people felt unwell or experienced symptoms characteristic of Covid-19 infection [24].

On 25 March New Zealand moved to a Level 4 ‘lockdown’, the highest level of alert and a National State of Emergency was declared [24]. At this alert level people were instructed to stay at home except for essential personal movement such as health or essential shopping, safe recreational activity is allowed in the local area, and travel is severely limited. All gatherings are cancelled, and all public venues are closed. Businesses are closed except for essential services (for example, supermarkets, pharmacies, health clinics, petrol stations and lifeline organisations). All educational facilities are closed [24].

As the spread of the virus slowed and stopped, the country progressively moved to lower alert levels: Level 3 towards the end of April and Level 2 in early May. Alert Level 1 was re-introduced on 8 June because community transmission had halted and there were no active cases in the country outside the Managed Isolation and Quarantine facilities (MIQ) specifically established to quarantine all incoming travellers to New Zealand for 14 days after arrival. If a traveller tested positive for Covid-19 at any time during the 14 days, they were moved to another quarantine facility for people with Covid-19 [24].

However, on 11August 2020 four new cases were detected in Auckland. Auckland returned the next day to Alert Level 3 with the rest of the country at Alert Level 2. Auckland remained at Alert Level 3 until 30 August, when it moved to Level 2, with additional restrictions on travel and the size of gatherings. The rest of the country remained at the standard Alert Level 2 until 21 September, when the alert level was downgraded to Level 1. The extra restrictions on Auckland residents were relaxed on 21 September and they returned to Alert Level 1 on 7 October 2020 [24].

## Materials and methods

A questionnaire seeking information from the public on their beliefs about, attitudes towards, and willingness to wear face masks, self-isolate and be tested for Covid-19 was designed based on the I_3_ Compliance Framework [6]. Involvement was measured using a condensed version of the Laurent and Kapferer [27] involvement scale developed by Kaine [28], with respondents rating two statements on each of the five components of involvement (functional, experiential, identity-based, risk-based, and consequence-based). Attitudes were measured using a simple, evaluative scale, while the strength of respondents’ attitudes, which were expected to vary depending on the strength of their involvement, was measured using an ipsative scale based on Olsen [29].

A series of questions was formulated to elicit respondents’ beliefs about Covid-19, eliminating Covid-19, wearing face masks, self-isolating and getting tested for Covid-19. Finally, information was sought on the demographic characteristics of respondents, including age, education, and ethnicity, and whether they wore masks, would self-isolate and had been tested for Covid-19.^2^

The ordering of the statements in the involvement, attitude and belief scales was randomised to avoid bias in responses. Respondents indicated their agreement with statements in all the involvement, attitude and belief scales using a five-point rating, ranging from strongly disagree (1) to strongly agree (5). The questionnaire is reproduced in S1.

Participation in the survey was voluntary, respondents could leave the survey at any time, and all survey questions were optional and could be skipped. The research approach was reviewed and approved by the Manaaki Whenua – Landcare Research’s social ethics process (application no. 2021/10 NK) which is based on the New Zealand Association of Social Science Research code of ethics.

The questionnaire was piloted with a small random sample (n=30), and subsequently completed by a large random sample (n=1001), of Auckland residents who were members of a large-scale, commercial consumer internet panel. Auckland was chosen for the survey because it is New Zealand’s largest city and is the mostly likely place for new community transmission to occur given the number of MIQ facilities and frontline border workers in the city.

Panel members receive reward points (which are redeemable for products and services) for completing surveys. An internet link to the questionnaire was distributed to a random sample of members of the panel subject to the constraint that they resided in Auckland and were not minors. The survey was conducted over two weeks from 7 September to 22 September 2020. Most of this time Auckland residents were under Alert Level 2, which meant they were expected to maintain social distancing when outside their homes and to wear masks in public places. They were also expected to keep track of their movements and to self-isolate and seek testing for Covid-19 if they felt unwell and experienced symptoms associated with Covid-19.

Given these circumstances, and given the pandemic had been receiving widespread coverage by mainstream and social media in New Zealand since February 2020, it seems reasonable to suppose that virtually all Auckland residents were aware of Covid-19 at the time we conducted our survey and most, if not all, were also aware of the government claims of the social desirability of wearing face masks and social distancing when out in public, of self-isolating if feeling unwell and getting tested for Covid-19. While awareness of the existence of Covid-19 is a prerequisite for involvement with it, awareness does not necessarily entail involvement. Widespread awareness of Covid-19 simply creates the potential for widespread involvement. The extent to which that potential is realised depends on respondents’ beliefs about how Covid-19 could affect the achievement of their functional, experiential and self-identity needs.

Awareness among Auckland residents of the social desirability of wearing face masks and social distancing when out in public, and of self-isolating if feeling unwell and getting tested for Covid-19, creates the strong possibility that self-reported past or likely future compliance with these preventive measures may be over-reported. Daoust et al. [30,31], for example, found this was the consistently case with respect to self-reported behaviour in the context of Covid-19 across a dozen countries, reporting compliance inflation of around 10% as a result of the design of survey questions [31]. Daoust et al. [31] found that this bias appeared to be consistent across gender, age, and education categories, which means the bias does not affect inferential comparisons made across these categories. However, the influence of bias in self-reporting socially desirable behaviours (or opinions) in the context of the categories in the I_3_ Framework is less clear (see the Discussion for more on this particular matter).

The focus of our survey is on the determinants of behavioural intentions and cognitive attention to messages. Where self-reported behaviour was sought in our survey, it was in the context of prior questions that explored impediments to, and specific aspects of, compliance and allowed a range of degrees, or frequency, of compliance rather than binary responses [30]. To this extent, the questioning effects that might evoke compliance bias in responses seem intrinsically weaker in this study compared to those reported in Daoust et al. [30,31].

As well, the online character of the survey reduces, if not removes [31], possible interviewer – based effects favouring over-reporting of compliance.

Of the 1,001 completed responses (giving a 99% confidence interval that population values are within 5% of the survey values), 53% were from women and 47% were from men. The age distribution aligned with the distribution for Auckland in the 2018 New Zealand Census, but Māori and Pacific Island residents were under-represented in the sample while European New Zealand residents were over-represented. Very-low-income households (<$20,000) and very-high-income households (>$100,000) were under-represented in the sample, while low-, middle- and high-income households were over-represented. Residents with secondary or certificate qualifications were substantially under-represented in the sample, while residents with graduate and postgraduate qualifications were substantially over-represented (see Appendix A).

Involvement scores were computed for each respondent as the simple arithmetic average of their agreement ratings for the 10 statements in the involvement scales. Attitudes scores were computed as the simple arithmetic average of their agreement ratings for the five statements in the attitude scales. Respondents were classified into belief segments based on their agreement ratings with the set of relevant belief statements using Ward’s method, with squared Euclidean distance as the measure of dissimilarity [32]. Note that, for all belief, involvement and evaluative attitudinal statements, respondents were instructed to indicate their agreement with a statement using a five-point rating scale, from strongly disagree (1) to strongly agree (5). Statistical analyses were conducted using the ‘cluster’ and ‘regression’ commands in SPSS [33].

## Results

### General

As shown in Table 1, all of the involvement and attitudinal scales with respect to eliminating Covid-19, wearing face masks, self-isolating when unwell and getting tested for Covid-19 exhibited satisfactory reliabilities [34]. Respondents’ involvement with the policy outcome (eliminating Covid-19) and wearing face masks, self-isolating when unwell, and getting tested for Covid-19 (the policy measures) are summarised in Tables 2, 3 and 4, respectively. Most respondents exhibited moderate-to-high involvement with eliminating Covid-19 from New Zealand and with the three policy measures (see [35] for more detail). However, a substantial minority of respondents exhibited low-to-mild involvement with wearing face masks and getting tested for Covid-19. This result suggests a minority of Auckland residents may inadvertently fail to comply with government measures intended to prevent the spread of Covid-19 in the community.

**Table 1.** Reliability of involvement and attitude scales

|  | Reliability<sup>1</sup> |
| --- | --- |
| Involvement with eliminating Covid-19 | 0.847 |
| Involvement with wearing face masks | 0.852 |
| Attitude towards wearing face masks | 0.854 |
| Involvement with self-isolating when unwell | 0.707 |
| Attitude towards self-isolating when unwell | 0.795 |
| Involvement with getting tested for Covid-19 | 0.821 |
| Attitude towards getting tested for Covid-19 | 0.849 |

Notes: <sup>1</sup> Values are Cronbach's alpha [34].

**Table 2.** I_3_ mapping for wearing face masks

| Quadrant | Favourable attitude | Unsure | Unfavourable attitude | Total |
| --- | --- | --- | --- | --- |
| Quadrant 1 | 3 | 4 | 2 | 9 |
| Quadrant 2 | 7 | 9 | - | 16 |
| Quadrant 3 | 59 | 11 | 3 | 73 |
| Quadrant 4 | 1 | - | - | 2 |
| Total | 70 | 24 | 5 | 100 |

Notes: Values are percentages of the sample.

**Table 3.** I_3_ mapping for self-isolating

| Quadrant | Favourable attitude | Unsure | Unfavourable attitude | Total |
| --- | --- | --- | --- | --- |
| Quadrant 1 | 1 | 1 | - | 2 |
| Quadrant 2 | 5 | 1 | 1 | 7 |
| Quadrant 3 | 74 | 6 | 5 | 85 |
| Quadrant 4 | 4 | 1 | 1 | 6 |
| Total | 84 | 9 | 7 | 100 |

Notes: Values are percentages of the sample.

**Table 4.** I_3_ mapping for testing

| Quadrant | Favourable attitude | Unsure | Unfavourable attitude | Total |
| --- | --- | --- | --- | --- |
| Quadrant 1 | 3 | 4 | - | 7 |
| Quadrant 2 | 14 | 6 | 1 | 21 |
| Quadrant 3 | 59 | 9 | 2 | 70 |
| Quadrant 4 | 1 | 1 | - | 2 |
| Total | 77 | 20 | 3 |  |

Notes: Values are percentages of the sample.

Most respondents exhibited a strongly favourable attitude, as measured by the ipsative attitude scale, towards mask wearing, self-isolating and getting tested. These results suggest a small minority of Auckland residents would deliberately choose not to comply with government measures intended to prevent the spread of Covid-19 in the community. These results and their implications for government policy are discussed in detail by Kaine [35].

The predominantly moderate-to-high involvement with, and favourable attitudes towards, measures (such as wearing face masks) to eliminate Covid-19 from New Zealand of most respondents is consistent with the high level of community endorsement of the New Zealand government’s approach to managing Covid-19 [36].

### Belief segments

Respondents’ beliefs were investigated because they can provide insights to guide the design of policies that, by modifying the beliefs and attitudes that underly compliance, seek to influence compliance. Using respondent agreement ratings for the relevant set of belief statements, respondents were classified into belief segments with respect to the nature of Covid-19, and the advantages and disadvantages of eliminating Covid-19 (the policy outcome), wearing face masks, self-isolating when unwell, and testing (the policy measures). Respondents were classified into segments using Ward’s method, with squared Euclidean distance as the measure of dissimilarity [32].

The number of segments was chosen based on the relative change in fusion coefficients, ease of interpreting the segments, and a desire to keep the number of segments as small as possible [32]. The segments, and their belief characteristics, are summarised below.

Beliefs about Covid-19, eliminating Covid-19, wearing masks, self-isolating when unwell and getting tested for Covid-19 were associated, to some extent, with demographic characteristics such as age, education, income, and ethnicity (see [35] for details).

#### Belief segments for Covid-19

Respondents were classified into five belief segments with respect to Covid-19 (Table 5). Most respondents had beliefs that align with accepted scientific facts. These respondents were classified as ‘Covid-19 convinced’ (41%) and ‘Covid-19 moderates’ (25%), the difference between these two segments being the intensity of their beliefs. The ‘Covid-19 safe healthy’ (9%) had beliefs that mostly align with accepted scientific facts, but these respondents believed Covid-19 only posed a danger to the elderly and people with health problems. A fourth segment, the ‘Covid-19 ambivalents’ (15%), consisted of respondents who were unsure about what to believe about Covid-19.

**Table 5.** Belief segments for Covid-19

| Statement | Covid-19<br>convinced (41%) | Covid-19<br>moderates<br>(25%) | Covid-19<br>ambivalents<br>(15%) | Covid-19 safe<br>healthy<br>(9%) | Covid-19<br>sceptics<br>(10%) |
| --- | --- | --- | --- | --- | --- |
| Coughing and sneezing spreads Covid-19 | 4.74 | 3.82<sup>a</sup> | 4.20<sup>a, b</sup> | 4.55<sup>b</sup> | 3.75<sup>a, c, d</sup> |
| Covid-19 spreads from surfaces touched by infected people | 4.15 | 3.78<sup>a</sup> | 3.97 | 4.11 | 3.57<sup>a, c, d</sup> |
| Covid-19 is only a danger to the elderly and people with health problems | 1.34 | 1.8<sup>a</sup> | 2.91<sup>a, b</sup> | 4.29<sup>a, b, c</sup> | 3.97<sup>a, b, c</sup> |
| You are immune to re-infection once you have had Covid-19 | 1.97 | 2.29<sup>a</sup> | 2.79<sup>a, b</sup> | 2.24<sup>c</sup> | 3.57<sup>a, b, c, d</sup> |
| Children cannot catch Covid-19 | 1.13 | 1.67<sup>a</sup> | 2.22<sup>a, b</sup> | 1.36<sup>b, c</sup> | 3.07<sup>a, b, c, d</sup> |
| Children are perfectly safe from Covid-19 | 1.25 | 1.70<sup>a</sup> | 2.17<sup>a, b</sup> | 1.47<sup>c</sup> | 3.43<sup>a, b, c, d</sup> |
| You cannot catch the virus from people without symptoms | 1.34 | 1.80<sup>a</sup> | 2.27<sup>a, b</sup> | 1.53<sup>c</sup> | 3.58<sup>a, b, c, d</sup> |
| Covid-19 is a hoax | 1.07 | 1.55<sup>a</sup> | 2.14<sup>a, b</sup> | 1.31 | 3.55<sup>a, b, c, d</sup> |
| Fears about Covid-19 are exaggerated | 1.55 | 2.24<sup>a</sup> | 3.31<sup>a, b</sup> | 2.28<sup>a, c</sup> | 3.78<sup>a, b, c, d</sup> |
| Covid-19 is no worse than the seasonal flu | 1.32 | 2.23<sup>a</sup> | 3.33<sup>a, b</sup> | 1.71<sup>a, b, c</sup> | 3.65<sup>a, b, d</sup> |
| Covid-19 is man-made | 2.24 | 2.57<sup>a</sup> | 3.08<sup>a, b</sup> | 2.62 | 3.84<sup>a, b, c, d</sup> |
| Covid-19 comes from bats | 2.97 | 2.95 | 2.74 | 2.96 | 3.48<sup>a, b, c, d</sup> |

Notes: Values are mean agreement ratings. Ratings ranged from a minimum of 1 (strongly disagree) to a maximum of 5 (strongly agree).
Differences in mean agreement ratings between segments tested using Tukey's HSD [37], ( $p < 0.01$ )
<sup>a</sup> Mean differs from mean for the 'convinced' segment
<sup>b</sup> Mean differs from mean for the 'moderates' segment
<sup>c</sup> Mean differs from the mean for the 'ambivalents' segment
<sup>d</sup> Mean differs from the mean for the 'safe healthy' segment

A small segment of respondents, the ‘Covid-19 sceptics’ (10%) believed, variously, that Covid-19 was a hoax, was no worse than the seasonal flu, and that fears about Covid-19 are exaggerated.

#### Belief segments for eliminating Covid-19

Respondents were classified into four belief segments with respect to eliminating Covid-19 (Table 6). Most respondents had beliefs that align with seeking to eliminate Covid-19 from New Zealand. These respondents were classified as ‘elimination enthusiasts’ (23%) and ‘elimination moderates’ (40%), the difference between these two segments being the intensity of their beliefs. Another segment of respondents, the ‘vaccination hopefuls’ (27%), agreed with trying to eliminate Covid-19 but were less sure that Covid-19 could be kept out of New Zealand indefinitely. They believed we must live with Covid-19 until a vaccine is available. A fourth segment, the ‘elimination sceptics’ (10%), consisted of respondents who believed we cannot eliminate Covid-19 indefinitely and we should try to build herd immunity.

**Table 6.** Belief segments for eliminating Covid-19

| Statement | Elimination<br>enthusiasts (23%) | Elimination<br>moderates (40%) | Vaccine<br>hopefuls (27%) | Elimination<br>sceptics (10%) |
| --- | --- | --- | --- | --- |
| We need to eliminate Covid-19 to save lives | 4.63 | 4.10 <sup>a</sup> | 3.56 <sup>a, b</sup> | 3.2 <sup>a, b, c</sup> |
| We should just live with it until we have a vaccine | 1.67 | 2.64 <sup>a</sup> | 3.61 <sup>a, b</sup> | 4.3 <sup>a, b, c</sup> |
| It would be better to let it spread and build herd immunity | 1.38 | 1.84 <sup>a</sup> | 2.57 <sup>a, b</sup> | 4.24 <sup>a, b, c</sup> |
| There is no point trying to eliminate Covid-19 because it is a virus and will keep changing | 1.7 | 2.56 <sup>a</sup> | 3.3 <sup>a, b</sup> | 4.38 <sup>a, b, c</sup> |
| Covid-19 is everywhere in the world so there is no way we can keep it out | 1.75 | 3.05 <sup>a</sup> | 4.00 <sup>a, b</sup> | 4.44 <sup>a, b, c</sup> |

Notes: Values are mean agreement ratings. Ratings ranged from a minimum of 1 (strongly disagree) to a maximum of 5 (strongly agree).
Differences in mean agreement ratings between segments tested using Tukey's HSD test [37] (p<0.01)
<sup>a</sup> Mean differs from mean for the 'enthusiasts' segment
<sup>b</sup> Mean differs from mean for the 'moderates' segment
<sup>c</sup> Mean differs from the mean for the 'hopefuls' segment

#### Wearing face masks and mask belief segments

Respondents were classified into four belief segments with respect to wearing face masks (Table 7). Most respondents believed that wearing face masks was effective in helping eliminate Covid-19 from New Zealand. These respondents were classified as ‘mask enthusiasts’ (45%) and ‘mask moderates’ (21%), the difference between these two segments being the intensity of their beliefs.

**Table 7.** Belief segments for wearing masks^1^

| Statement | Mask enthusiasts<br>(45%) | Mask moderates<br>(21%) | Mask ambivalents<br>(27%) | Mask sceptics<br>(7%) |
| --- | --- | --- | --- | --- |
| Masks are effective | 4.48 | 3.78<sup>a</sup> | 3.34<sup>a, b</sup> | 3.24<sup>a, b</sup> |
| Wearing masks should be compulsory | 4.04 | 3.53<sup>a</sup> | 2.8<sup>a, b</sup> | 2.90<sup>a, b</sup> |
| Wearing a mask sets a good example to others | 4.52 | 4.09<sup>a</sup> | 3.41<sup>a, b</sup> | 3.34<sup>a, b</sup> |
| You should only have to wear a mask if you feel unwell | 1.5 | 1.96<sup>a</sup> | 3.39<sup>a, b</sup> | 4.24<sup>a, b, c</sup> |
| You should only have to wear a mask if you are old or have a health problem | 1.5 | 1.78<sup>a</sup> | 3.05<sup>a, b</sup> | 4.06<sup>a, b, c</sup> |
| People who wear masks are over-reacting | 1.39 | 1.99<sup>a</sup> | 2.85<sup>a, b</sup> | 4.28<sup>a, b, c</sup> |
| Masks are not practical | 1.53 | 2.22<sup>a</sup> | 3.10<sup>a, b</sup> | 4.26<sup>a, b, c</sup> |
| Home-made masks are a waste of time and effort | 1.77 | 2.50<sup>a</sup> | 2.96<sup>a, b</sup> | 4.56<sup>a, b, c</sup> |
| Masks are just too uncomfortable | 2.18 | 2.99<sup>a</sup> | 3.41<sup>a, b</sup> | 4.14<sup>a, b, c</sup> |
| Masks we can buy are not worth bothering with | 1.93 | 2.53<sup>a</sup> | 3.13<sup>a, b</sup> | 4.38<sup>a, b, c</sup> |
| Masks are too difficult if you wear glasses | 2.36 | 3.35<sup>a</sup> | 3.49<sup>a</sup> | 4.16<sup>a, b, c</sup> |
| Masks are not much help without gloves | 2.02 | 2.66<sup>a</sup> | 3.10<sup>a, b</sup> | 3.70<sup>a, b, c</sup> |
| Masks on their own are not much help | 2.13 | 3.13<sup>a</sup> | 3.28<sup>a</sup> | 4.24<sup>a, b, c</sup> |
| Masks are not much help because they are not worn properly | 2.43 | 2.98<sup>a</sup> | 3.49<sup>a, b</sup> | 4.42<sup>a, b, c</sup> |

Notes: Values are mean agreement ratings. Ratings ranged from a minimum of 1 (strongly disagree) to a maximum of 5 (strongly agree).
Differences in mean agreement ratings between segments tested using Tukey’s HSD test [37] (p<0.01)
<sup>a</sup> Mean differs from mean for the ‘enthusiasts’ segment
<sup>b</sup> Mean differs from mean for the ‘moderates’ segment
<sup>c</sup> Mean differs from the mean for the ‘ambivalents’ segment

Another segment of respondents, the ‘mask ambivalents’ (27%), agreed masks could be effective but were less sure about the need to wear masks if you were young and healthy, and the usefulness of masks on their own, and doubted the effectiveness of masks that were home-made or available for purchase by the public. A fourth segment consisted of respondents, the ‘mask sceptics’ (7%), who were not convinced masks were effective. These respondents believed you were over-reacting if you wore a mask unless you were elderly or had a health problem. They also believed that masks were of limited usefulness on their own and doubted the quality of masks that were home-made or could be purchased by the public.

#### Self-isolating when unwell and self-isolation belief segments

Respondents were classified into three belief segments with respect to self-isolating (Table 8). Most respondents believed that self-isolating, if you felt unwell or had any of the symptoms associated with Covid-19, was effective in helping eliminate Covid-19 from New Zealand. These respondents were classified as ‘self-isolation enthusiasts’ (60%). Another large group of respondents, the ‘self-isolation ambivalents’ (29%), also believed that self-isolating was effective in helping eliminate Covid-19 but were unsure about the practicalities of it.

**Table 8.**
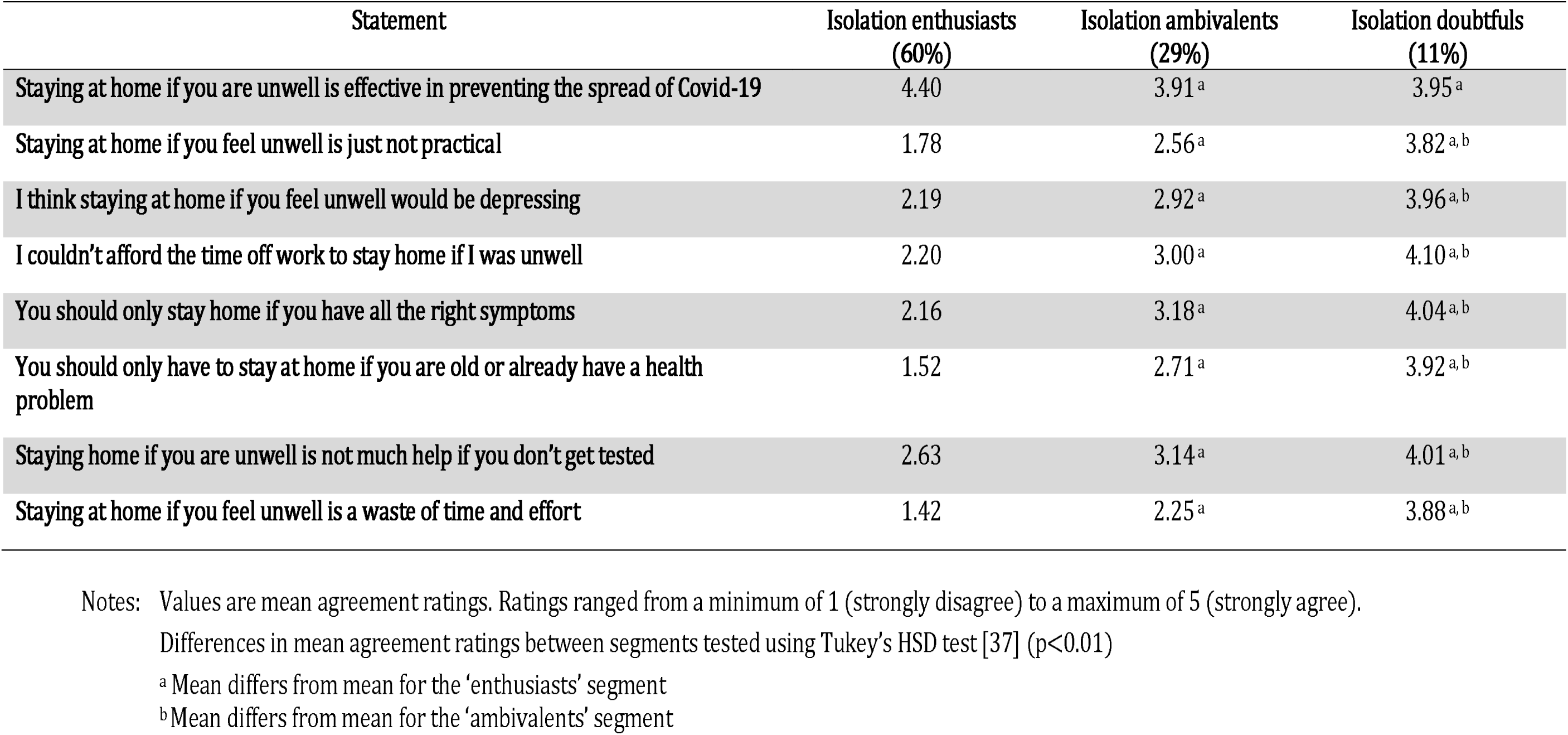
Belief segments for self-isolating

A third, smaller, segment consisted of the ‘self-isolation doubtfuls’ (11%), who believed self-isolating was effective in preventing the spread of Covid-19 but did not believe it was practical and would most likely be a waste of their time. These respondents believed that they could not afford the time off work to self-isolate, and you should only have to self-isolate if you were old, already had a health problem, or had all the right symptoms. They also believed staying at home if you were unwell was not much help if you didn’t get tested.

#### Covid-19 testing and testing belief segments

Respondents were classified into four belief segments with respect to testing for Covid-19 (Table 9). Nearly all respondents believed that testing for Covid-19 was effective in helping eliminate Covid-19 from New Zealand. However, respondents differed in their beliefs about the efficacy of tests and who should be tested.

**Table 9.**
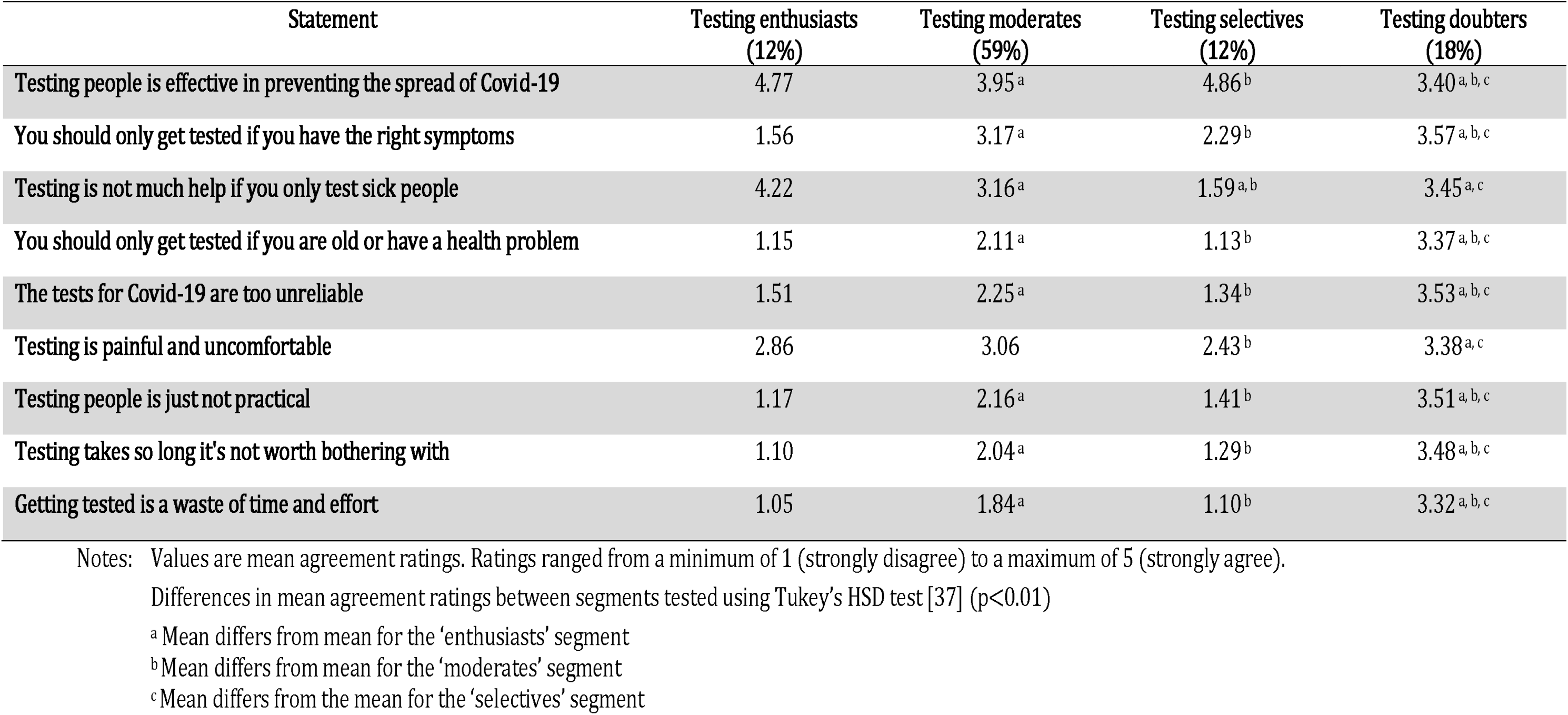
Belief segments for Covid-19 testing

| Statement | Testing enthusiasts<br>(12%) | Testing moderates<br>(59%) | Testing selectives<br>(12%) | Testing doubters<br>(18%) |
| --- | --- | --- | --- | --- |
| Testing people is effective in preventing the spread of Covid-19 | 4.77 | 3.95<sup>a</sup> | 4.86<sup>b</sup> | 3.40<sup>a, b, c</sup> |
| You should only get tested if you have the right symptoms | 1.56 | 3.17<sup>a</sup> | 2.29<sup>b</sup> | 3.57<sup>a, b, c</sup> |
| Testing is not much help if you only test sick people | 4.22 | 3.16<sup>a</sup> | 1.59<sup>a, b</sup> | 3.45<sup>a, c</sup> |
| You should only get tested if you are old or have a health problem | 1.15 | 2.11<sup>a</sup> | 1.13<sup>b</sup> | 3.37<sup>a, b, c</sup> |
| The tests for Covid-19 are too unreliable | 1.51 | 2.25<sup>a</sup> | 1.34<sup>b</sup> | 3.53<sup>a, b, c</sup> |
| Testing is painful and uncomfortable | 2.86 | 3.06 | 2.43<sup>b</sup> | 3.38<sup>a, c</sup> |
| Testing people is just not practical | 1.17 | 2.16<sup>a</sup> | 1.41<sup>b</sup> | 3.51<sup>a, b, c</sup> |
| Testing takes so long it's not worth bothering with | 1.10 | 2.04<sup>a</sup> | 1.29<sup>b</sup> | 3.48<sup>a, b, c</sup> |
| Getting tested is a waste of time and effort | 1.05 | 1.84<sup>a</sup> | 1.10<sup>b</sup> | 3.32<sup>a, b, c</sup> |

Notes: Values are mean agreement ratings. Ratings ranged from a minimum of 1 (strongly disagree) to a maximum of 5 (strongly agree).
Differences in mean agreement ratings between segments tested using Tukey's HSD test [37] ( $p < 0.01$ )
<sup>a</sup> Mean differs from mean for the 'enthusiasts' segment
<sup>b</sup> Mean differs from mean for the 'moderates' segment
<sup>c</sup> Mean differs from the mean for the 'selectives' segment

Most respondents believed that testing was practical and reliable, and should include the healthy as well as the elderly, people with health problems or people with Covid-19 symptoms. These respondents were classified as ‘testing enthusiasts’ (12%) and ‘testing moderates’ (59%), the difference between these two segments being the intensity of their beliefs.

Another segment of respondents, the ‘testing selectives’ (12%), were like the ‘testing enthusiasts’ in believing that testing was practical and reliable, but they believed testing could be limited to sick people. A fourth segment, the ‘testing doubters’ (18%), consisted of respondents who believed testing was effective in preventing the spread of Covid-19, but did not believe it was practical or reliable, and that testing should be limited to the elderly, people with health problems and people with Covid-19 symptoms.

Compared to the other testing segments, a relatively high proportion of ‘testing enthusiasts’ indicated they had been tested for Covid-19. There were no differences among the segments in the proportion of respondents who had been tested in each segment and who had felt unwell when they were tested. Assuming the probability of exposure to Covid-19 and the probability of ‘feeling unwell’ is similar across the segments, one explanation for this result is that respondents in the ‘testing enthusiasts’ segment are more likely than those in other segments to seek testing, whether they are well or unwell.

While a relatively high proportion of Māori and Pacific Islander respondents in the sample had been tested for Covid-19, a relatively low proportion of respondents from other ethnic groups, including European New Zealanders, had been tested. A relatively high proportion of respondents who were European New Zealanders who had been tested were unwell at the time of testing, while a relatively low proportion of respondents who were Māori or Pacific Islanders were unwell when tested.

There were no other socio-demographic differences between respondents in the sample who had been tested for Covid-19 and those who had not [35].

### Predicting compliance

The purpose of this analysis was to quantify the effect of beliefs, attitudes, and involvement on respondents’ propensity to comply with wearing face masks in public, self-isolating and being tested for Covid-19. In other words, we wanted to estimate, separately, the influence of involvement (as a measure of the strength of individuals’ motivation) on compliance and the influence of beliefs and attitudes in the context of eliminating Covid-19 from New Zealand by wearing masks, self-isolating when unwell and being tested.

We hypothesised that the propensity to wear face masks, self-isolate when unwell and get tested for Covid-19 was a function of involvement and attitude, as outlined by Kaine et al. [6]. We also hypothesised that the marginal effect of an increase in involvement with the outcome (eliminating Covid-19) would decrease with higher levels of involvement with the measure (e.g. wearing face masks). The same would apply with respect to the marginal effect of an increase in involvement with the measure (e.g. wearing face masks), which would decrease with higher levels of involvement with the outcome (eliminating Covid-19). In addition, we hypothesised that attitudes towards mask wearing, self-isolating when unwell and testing were a function of beliefs about Covid-19, eliminating Covid-19, mask wearing, self-isolating and testing.

To summarise, two sets of regressions were estimated. One set had propensity to comply as the dependent variable (e.g. wear face masks), with involvement with the outcome (eliminating Covid-19), involvement with the measure (e.g. wearing masks), the interaction between the involvement with the outcome and the measure, and attitudes towards the measure as the explanatory variables.

The second set had attitudes towards the measures (wearing face masks, self-isolating and testing) as the dependent variables, with beliefs about Covid-19, eliminating Covid-19, and beliefs about the measures (mask wearing, self-isolating when unwell and testing respectively) as the explanatory variables.

Respondents’ propensity to wear face masks was obtained by asking them if they had worn a face mask when out in public the previous week and if they had to go out to work the previous week. Respondents answered both questions using a five-point scale ranging from ‘always’ to ‘never’. Their propensity to self-isolate was obtained by asking them, ‘Thinking about the next few days, would you stay home if you were unwell or had any of the following symptoms: a dry cough, fever, loss of sense of smell, loss of sense of taste, shortness of breath or difficulty breathing?’. We also asked, ‘If you were advised to do so by a health care professional or public health authority, would you self-isolate for 14 days?’. Both questions were answered using a five-point scale ranging from ‘definitely’ to ‘definitely not’.

Regarding testing, the propensity to be tested will depend on their perceptions of the risk of exposure to Covid-19 and whether they had experienced symptoms associated with Covid-19, as well as their involvement with, and attitudes towards, testing. We had data indicating whether respondents had been tested for Covid-19 and, if they had, whether they were feeling unwell at the time. However, we did not have data indicating whether any respondents had felt unwell but had not sought Covid-19 testing. Nor did we have data on respondent’s perceptions of exposure to Covid-19 such as their proximity to sources of outbreaks. Consequently we were unable to estimate satisfactory regressions for being tested.

Dummy variables were created representing respondents’ membership of belief segments with respect to Covid-19, eliminating Covid-19, mask wearing, self-isolating when unwell, and being tested. In each instance, the ‘Covid-19 convinced’ or ‘enthusiast’ segments were treated as the benchmark. Attitudes towards the policy measures were included using the evaluative scale described earlier.

The explanatory power of the regressions, and the resulting parameter estimates, are reported in Tables 10 and 11. The compliance regressions were statistically significant and, for cross-sectional data, a substantial proportion of the variance in respondents’ compliance was explained by their involvement and attitudes (apart from the regression for staying at home if feeling unwell).

**Table 10.**
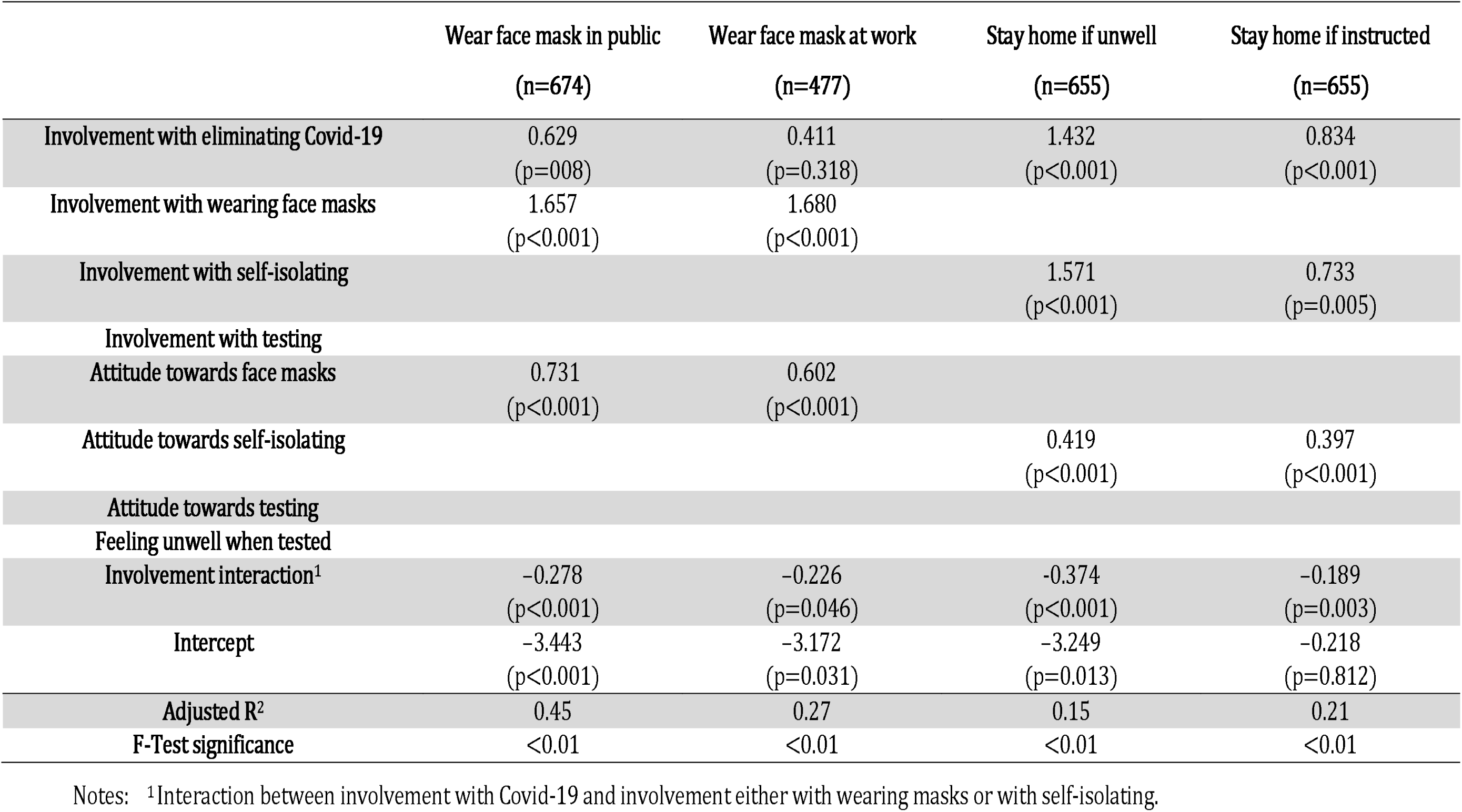
Parameter estimates for propensity to wear face masks and self-isolate

|  | Wear face mask in public<br>(n=674) | Wear face mask at work<br>(n=477) | Stay home if unwell<br>(n=655) | Stay home if instructed<br>(n=655) |
| --- | --- | --- | --- | --- |
| Involvement with eliminating Covid-19 | 0.629<br>(p=008) | 0.411<br>(p=0.318) | 1.432<br>(p<0.001) | 0.834<br>(p<0.001) |
| Involvement with wearing face masks | 1.657<br>(p<0.001) | 1.680<br>(p<0.001) |  |  |
| Involvement with self-isolating |  |  | 1.571<br>(p<0.001) | 0.733<br>(p=0.005) |
| Involvement with testing |  |  |  |  |
| Attitude towards face masks | 0.731<br>(p<0.001) | 0.602<br>(p<0.001) |  |  |
| Attitude towards self-isolating |  |  | 0.419<br>(p<0.001) | 0.397<br>(p<0.001) |
| Attitude towards testing |  |  |  |  |
| Feeling unwell when tested |  |  |  |  |
| Involvement interaction<sup>1</sup> | -0.278<br>(p<0.001) | -0.226<br>(p=0.046) | -0.374<br>(p<0.001) | -0.189<br>(p=0.003) |
| Intercept | -3.443<br>(p<0.001) | -3.172<br>(p=0.031) | -3.249<br>(p=0.013) | -0.218<br>(p=0.812) |
| Adjusted R<sup>2</sup> | 0.45 | 0.27 | 0.15 | 0.21 |
| F-Test significance | <0.01 | <0.01 | <0.01 | <0.01 |

Notes: <sup>1</sup>Interaction between involvement with Covid-19 and involvement either with wearing masks or with self-isolating.

**Table 11.** Parameter estimates for attitude towards wearing face masks, self-isolating and seeking testing

|  | Attitude towards face<br>masks<br>(n=682) | Attitude towards self-isolating<br>(n=655) | Attitude towards<br>testing<br>(n=662) |
| --- | --- | --- | --- |
| Covid-19 moderate | -0.194<br>(p<0.001) | -0.111<br>(p=0.035) | -0.206<br>(P<0.001) |
| Covid-19 ambivalent | -0.307<br>(p<0.001) | -0.153<br>(p=0.028) | -0.308<br>(p<0.001) |
| Covid-19 healthy | -0.171<br>(p=0.031) | -0.064<br>(p=0.452) | -0.034<br>(p=0.655) |
| Covid-19 sceptic | -0.328<br>(p<0.001) | -0.378<br>(p<0.001) | -0.309<br>(P<0.001) |
| Elimination moderate | 0.049<br>(p=0.389) | -0.035<br>(p=0.535) | -0.028<br>(p=0.603) |
| Elimination hopeful | -0.079<br>(p=0.228) | -0.049<br>(p=0.427) | -0.053<br>(p=0.378) |
| Elimination sceptic | 0.059<br>(p=0.539) | 0.101<br>(p=0.258) | 0.100<br>(p=0.255) |
| Mask moderate | -0.387<br>(p<0.001) |  |  |
| Mask ambivalent | -1.133<br>(P<0.001) |  |  |
| Mask sceptic | -1.532<br>(p<0.001) |  |  |
| Isolation ambivalent |  | -0.461<br>(p<0.001) |  |
| Isolation doubter |  | -0.845<br>(p<0.001) |  |
| Test selectives |  |  | 0.000<br>(p=0.992) |
| Test moderates |  |  | -0.556<br>(p<0.001) |
| Test doubters |  |  | -1.262<br>(p<0.001) |
| Intercept | 4.715<br>(p<0.001) | 4.727<br>(p<0.001) | 4.849<br>(p<0.001) |
| Adjusted R<sup>2</sup> | 0.55 | 0.30 | 0.45 |
| F-Test significance | <0.01 | <0.01 | <0.01 |

The attitudinal regressions were statistically significant and, for cross-sectional data, a substantial proportion of the variance in the attitudes of respondents was explained by their beliefs.

Willingness to wear face masks in public and at work was strongly and positively influenced by involvement with eliminating Covid-19 as well as involvement with, and attitudes towards, wearing face masks. Willingness to self-isolate when unwell was also strongly and positively influenced by involvement with eliminating Covid-19 as well as involvement with, and attitudes towards, self-isolating.

As hypothesised, the marginal impact of an increase in involvement with wearing face masks or self-isolating when unwell decreased with higher levels of involvement with eliminating Covid-19 (and vice versa) and illustrated in S3 with respect to wearing face masks. Overall, the results reported in Table 10 support the idea that involvement and attitudes have differential effects on compliance.

Respondents’ attitudes towards wearing face masks were influenced by their beliefs about Covid-19 and about the effectiveness of wearing face masks (see Table 11). Similarly, their attitudes towards self-isolating when unwell were influenced by their beliefs about Covid-19 together with their beliefs about the effectiveness of self-isolating. Lastly, their attitudes towards testing were influenced by their beliefs about Covid-19 together with their beliefs about the effectiveness of testing. Respondents’ beliefs about the effectiveness of eliminating Covid-19 as a strategy did not appear to have a significant influence on attitudes to any of the measures. In all instances the signs on the estimated parameters were consistent with expectations with attitudes becoming more and more unfavourable as respondents’ beliefs shifted towards scepticism as has been found in other studies [38,39].

## Discussion

Kaine et al. [6] proposed that the propensity of individuals to change their behaviour and comply with policy measure depends on the intensity of their involvement with the policy outcome and policy measure, as well as their attitude towards the policy measure. The results presented here largely support that proposition: they clearly indicate that involvement (how much an individual cares about a subject) influences the propensity to comply with measures to prevent the spread of Covid-19 in addition to their attitude towards those measures. The prosaic but important implication is that people may hold similar opinions or attitudes towards a protocol such as wearing face masks in public, but their propensity to do so may vary markedly depending on how involved they are with – how much they care about – preventing the spread of Covid-19 and wearing face masks.

Of course, the influence of involvement becomes unimportant in explaining differences in compliance if everyone has a similar level of involvement, as appears to be the case with self-isolating when unwell. In these circumstances, individual differences in attitudes and constraints on behaviour, such as the capacity to absorb salary losses, are the determinants of compliance.

Our findings have three important implications for promoting community compliance with measures intended to prevent the spread of Covid-19 in particular, and the design of policy measures generally. The first is that people may, inadvertently, fail to comply with a measure even though they may have favourable attitudes towards the policy outcome, simply because they are not paying attention. In circumstances where involvement is low, compliance (or non-compliance) is not a matter of deliberate choice. Consequently, authorities must consider carefully imposing blanket penalties for non-compliance because they run the risk of alienating people who would otherwise do the ‘right thing’.

In the context of measures to prevent the spread of Covid-19, this translates into ensuring that compliance requires as little effort and thought as practical, and is as stress-free as possible [40]; for example, by supplying face masks for free on public transport and other high-risk locations such as supermarkets [41], by ensuring testing is as convenient as possible, by minimising as far as practical the time spent travelling to testing centres and the time spent queuing for tests, and by offering limited compensation for those who are required to self-isolate because they test positive.

In these circumstances, where involvement is high, compliance (or non-compliance) is most likely to be a deliberate choice, the choice depending on one’s attitude towards the wearing of face masks, self-isolating when unwell, and testing. If attitudes are strongly unfavourable, then severe penalties or substantial inducements may be required to secure compliance, or compliance could be imposed (for example, compelling employers to report staff with Covid-19 symptoms to health authorities).

Clearly, differences in the level of involvement people have with the outcome of eliminating Covid-19 and the measures for preventing the spread of Covid-19 create an additional complication for authorities responsible for implementing the strategy. Tactics to promote compliance among those with low involvement will not influence the highly involved who are non-compliant, unless the latter can be rapidly and easily distinguished from the former.

The second implication is that people who have low involvement with the policy outcome and the policy measures may miss important promotional messages simply because they are not paying attention. In circumstances where involvement with a subject is low, sensitivity to promotional messages about the subject is low. Messages are not necessarily deliberately ignored; they simply fail to catch the attention of those with low involvement (they are not noticed).

In the context of Covid-19 measures, this means that people with low-to-mild involvement with eliminating Covid-19 and with the measures for preventing the spread of Covid-19 may fail to notice or properly process promotional messages about Covid-19 and the measures. They may, for example, be entirely unaware of lockdown rules (or even what level of lockdown is in play). This increases the risk that people with low involvement may inadvertently be non-compliant, especially if changes are made to lockdown rules, or new lockdown levels are introduced.

Relatedly, Gray et al., [42] found, as we did, that people’s willingness to wear face masks depended on their beliefs about the effectiveness of face masks in protecting them from infection. Consequently, they recommended developing and promoting to the public clear guidelines on wearing face masks and increasing promotional efforts dispelling myths about the efficacy of masks [42]. Such promotional efforts are unlikely to meet with success among people who have low-to-mild involvement with Covid-19 and with wearing face masks as they are unlikely to pay proper attention to such promotional messages unless they are designed in such a way as to capture their attention.

The attention of people with low involvement in a subject can be captured if messages about the subject can be linked to another matter that is involving for them. This requires identifying, for those not interested in Covid-19, themes that are involving for them, and that can be meaningfully linked to containing the spread of Covid-19. The sport metaphor ‘[We are] a team of five million’, employed in community messaging about Covid-19 by the New Zealand government, is one example, though this metaphor may not be universally appealing. Other examples may be framing messages about following Covid-19 measures in the context of protecting families and jobs [36].

The third implication concerns the intrinsic malleability of the beliefs and attitudes of people who have low involvement with a subject. Such people devote little time and effort to gathering information about the subject, evaluating that information, and forming beliefs about and attitudes towards the subject. This means their beliefs and attitudes may be unstable and can change rapidly.

With respect to preventing the spread of Covid-19, this raises the possibility that, on the one hand, the distribution of misinformation through social media may provoke changes in the beliefs and attitudes of people with low involvement in Covid-19 that are undesirable because they undermine compliance with Covid-19 measures [39,43]. Such misinformation may provide a self-serving rationale for failing to comply with measures that require an investment of time and effort. On the other hand, people with low involvement are unlikely to strongly endorse misinformation (unless it is framed within a context they find highly involving) and so are unlikely to be provoked into engaging in non-compliant behaviours that require an investment of time and effort (such as attending protest rallies).

These considerations suggest that government authorities must be careful to discriminate between audiences on social media in terms of involvement when it comes to investing resources in combating misinformation about Covid-19. Presumably, those with low involvement in eliminating Covid-19 will exhibit a lower intensity and pattern of engagement with misinformation on social media than those with high involvement.

The willingness of people to adopt behaviours to prevent the spread of Covid-19 such as wearing face masks, self-isolating and getting tested for Covid-19 has been the subject of numerous studies [41,44,45,46,47,48,49,50]. These studies have shown that willingness to adopt these behaviours does depend on peoples’ attitudes towards them, which in turn depend on their beliefs about the behaviours. Consequently, many of these studies recommend that the adoption of preventive behaviours can be improved through promotional efforts intended to change beliefs and attitudes. Our results have two important implications for such recommendations.

First, while it is undoubtedly true that changing attitudes can change behaviour, promotional efforts intended to change beliefs and attitudes about preventative behaviours are unlikely to meet with complete success unless most people have moderate-to-high involvement both with preventing the spread of Covid-19 and with preventative behaviours.

Second, the greater the proportion of people with low-to-mild involvement with preventing the spread of Covid-19 and with preventative behaviours, the more likely efforts to improve the ease and convenience of adopting preventative behaviours will be more effective in changing behaviour compared to promotional efforts aimed at changing peoples’ beliefs and attitudes. See West et al., [51] for a discussion about the range of interventions that might accompany promotion.

Our findings are subject to a several qualifications including the following. First, beliefs, attitudes, and behaviours regarding preventing the spread of Covid-19, the wearing of face masks, self-isolating when unwell and getting tested for Covid-19 may have changed over time as the pandemic has progressed. Second, as the survey sample was drawn from an internet panel there may be selection bias. While the nature and severity of this bias in relation to the beliefs, attitudes, and involvement we investigated is unknown, it does seem reasonable to suppose, ceteris paribus, that people with low-to-mild involvement may be under-represented in the sample.

Third, as the scales measuring wearing of face masks, willingness to self-isolate and being tested for Covid-19 were self-reported, our measurements of these behaviours may have been affected by social desirability bias [30]. As noted above, while Daoust et al. [31] found that social desirability bias appeared to be consistent across gender, age, and education categories, the potential for bias in self-reporting of socially desirable behaviours (or opinions) in the context of the categories in the I_3_ Framework is less clear. There may be correlation between quadrant membership and social desirability bias.

On one hand, those with low involvement in, say, eliminating Covid-19 and wearing face masks might be expected to exhibit (downward) social desirability bias in self-reporting behaviours because they are less motivated to actually engage in the behaviour. However, their lack of interest in eliminating Covid-19 and wearing face masks means they may be quite insensitive to the opinions of others about the behaviour, and so be less susceptible to social desirability bias.

On the other hand, those with high involvement in eliminating Covid-19 and wearing face masks might be expected to exhibit (upward) social desirability bias in self-reporting behaviours because they are motivated to actually engage in the behaviour. Their very interest in eliminating Covid-19 and wearing face masks may mean they are quite sensitive to the opinions of others about the behaviour (especially if expressing self-identity is an important source of involvement) and thus be more susceptible to social desirability bias. On this argument, self-reporting of socially desirable behaviours like wearing face masks might mean the actual association between involvement and engagement in such behaviours could be over-estimated. As noted earlier, our expectation is that the design and administration of the survey are such as to reduce the likely magnitude of social-desirability bias effects compared to previous findings. Clearly, the interaction between I_3_ quadrant membership and social desirability bias is an area requiring further study.

Fourth, the adoption of behaviours such as the wearing of face masks has been associated with a range of variables including perceptions of the perceived risk of infection, the local incidence rate of COVID-19 and feelings of stress in relation to COVID-19 (see [40] for example). We did not include these variables in our analysis, and, while the correlation between these variables and involvement is unknown, it is likely to be positive.

## Conclusions

Governments around the world are seeking to slow the spread of Covid-19 by implementing measures that encourage, or mandate, changes in people’s behaviour. These changes include the wearing of face masks, social distancing, and testing and self-isolating when unwell. The success of these measures depends on the commitment of individuals to change their behaviour accordingly. Understanding and predicting the motivation of individuals to change their behaviour is critical to assessing the likely effectiveness of these measures in slowing the spread of the virus.

Kaine et al. [6] hypothesised that the propensity of individuals to change their behaviour and comply with policy measures depends on the intensity of their involvement with, and their attitude towards, the measure. This is because cognitive effort is required to form a strongly held attitude, and such effort is only invested when the matter at hand is sufficiently important to the individual. They also hypothesised that the propensity of individuals to comply with policy measures also depends on their involvement with the policy outcome the measure addresses. An implication of these hypotheses is that individuals with similar attitudes will display varying degrees of compliance with policy measures depending on the intensity of their involvement with the policy outcome and the policy measure.

We tested these hypotheses, and their implication, using compliance with measures to prevent the spread of Covid-19 in New Zealand. Broadly speaking the hypotheses and their implication are supported by the results. The finding that compliance depends on involvement (motivation) as well as attitude has important implications for the design of policy measures intended to promote compliance. This finding also has important implications for the design of promotional programmes to communicate information to the community about policy measures intended to promote compliance.

With respect to preventing the spread of Covid-19 in New Zealand, the results highlight the importance of distinguishing unintentional non-compliance with respect to wearing face masks, self-isolating when unwell, and testing from deliberate non-compliance, and tailoring enforcement strategies appropriately. The results also highlight the difficulty of communicating effectively through mass media with those who have low involvement with preventing the spread of Covid-19, and the importance of distinguishing between those with low and high involvement in considering the possible effects on compliance of the dissemination of misinformation about Covid-19 through social media.

## Supporting information

S1 Questionnaire

## Data Availability

All data produced in the present study are available upon reasonable request to the authors

## Acknowledgements

We would sincerely like to thank the residents of Auckland who completed the questionnaire. Thanks also to our two anonymous referees for their time, patience, constructive advice, and suggestions and for bringing the potential implications of social desirability bias to our attention.

## Supporting information

**S1 File. Questionnaire**

**S2 File. Data**

**S3 Fig. S3 Fig**

**S4 File. Appendix A**

---

**Fig S3.**
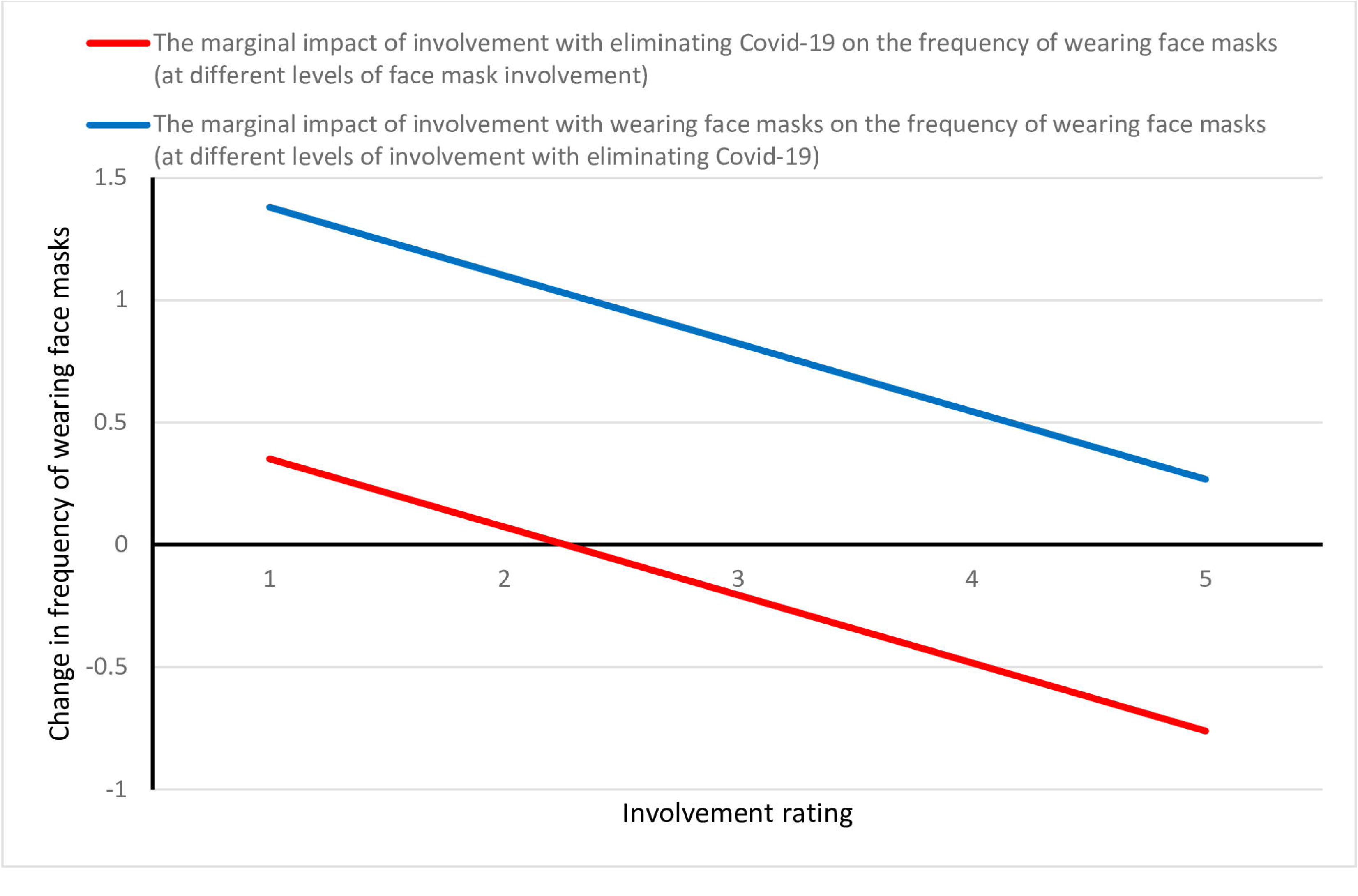
Changes in the marginal impact of increasing involvement with eliminating Covid-19 and wearing face masks on frequency or wearing face masks. The marginal impact of an increase in involvement with wearing face masks on frequency decreases with higher levels of involvement with eliminating Covid-19 (and vice versa). For example, the red line shows that an increase in involvement with eliminating Covid-19 has a decreasing impact on frequency of wearing face masks as involvement with wearing face masks increases. Involvement was rated on a scale from a minimum of 1 (low) to a maximum of 5 (high). Frequency was rated on a scale from a minimum of 1 (never) to a maximum of 5 (always).

## Footnotes

1 *I*nvolvement, *i*ssue and *i*ntervention, hence I_3_ Framework.

2 The ethnicity categories were Māori (the indigenous people of New Zealand), European New Zealander, Pacific Islander, Asian and Other.

## References

1. Czeisler ME, Tynan MA, Howard ME, Honeycutt S, Fulmer EB, Kidder DP, et al, Public attitudes, behaviors, and beliefs related to COVID-19, stay-at-home orders, nonessential business closures, and public health guidance – United States, New York City, and Los Angeles, May 5-12, 2020. Morbidity and Mortality Weekly Report. 2020; 69(24): 751–758. doi.org/10.15585/mmwr.mm6924e1

2. Hager E, Odetokun IA, Bolarinwa O, Zainab A, Okechukwu O, Al-Mustapha AI, Knowledge, attitude, and perceptions towards the 2019 Coronavirus Pandemic: A bi-national survey in Africa. PLoS ONE, 2020; 15(7): e0236918. doi.org/10.1371/journal.pone.0236918

3. Jarynowski A, Wójta-Kempa M, Płatek D, Czopek K, Attempt to understand public health relevant social dimensions of COVID-19 outbreak in Poland (5 April 2020). Available at SSRN: https://ssrn.com/abstract=3570609

4. Burby RJ, Paterson RG, Improving compliance with state environmental regulations. J. Policy Anal. Manage. 1993; 12: 753–772.

5. Daoust J.-F, 2020. Elderly people and responses to COVID-19 in 27 Countries. PLoS ONE 15(7): e0235590. https://doi.org/10.1371/journal.pone.0235590

6. Kaine G, Murdoch H, Lourey R, Bewsell D, A framework for understanding individual response to regulation. Food Policy 2010; 35: 531–537.

7. Kim Y, Conceptualizing health campaign strategies through the level of involvement. Corp. Commun. Int. J. 2003; 8: 255–267.

8. Zaichkowsky JL, Measuring the involvement construct. J. Consum. Res. 1985; 12: 341–352.

9. Ajzen I, Fishbein M, Attitude-behaviour relations: A theoretical analysis and review of empirical research. Psychol. Bull. 1977; 84: 888–918.

10. Kassarjian HH, Low involvement: A second look, in: Monroe KB. (Ed.), Advances in Consumer Research. Association for Consumer Research, 1981; 31–34.

11. Murdoch H, Bewsell D, Lourey R, Kaine G, Understanding people’s response to biosecurity regulation. Decision Making in Uncertain Times, 3rd National Conference on Risk Management. The New Zealand Society for Risk Management Inc, Auckland, 2006.

12. Chaffee SH, Roser C, Involvement and the consistency of knowledge, attitudes, and behaviours. Commun. Res. 1986; 13: 373–399.

13. Petty RE, Cacioppo JT, The Effects of Involvement on Responses to Argument Quantity and Quality: Central and Peripheral Routes to Persuasion. J. Pers. Soc. Psychol. 1984; 46: 69–81.

14. Thaler R.H, Sunstein CR, Nudge: improving decisions about health, wealth, and happiness. New Haven, CT: Yale University Press; 2008.

15. Gunningham N, Grabosky P, Sinclair D, Smart regulation: Designing environmental policy. Oxford University Press, New York; 1998.

16. Davies A, Kaine G, Lourey R, Understanding factors leading to non-compliance with effluent regulations by dairy farmers. Environment Waikato Technical Report 2007/37, Environment Waikato, Hamilton; 2007.

17. Carlough L, General deterrence of environmental violation: A peek into the mind of the regulated public. Oregon Department of Environmental Quality. 2003. Available at http://www.deq.state.or.us/programs/enforcement [accessed June 6 2008].

18. Kaine G, Tostovrsnik N, Landholders and the management of weeds: Blackberry and serrated tussock. Service Design Research Working Paper 03-11, Department of Primary Industries, Tatura, Victoria; 2011.

19. Lourey R, Kaine G, Davies A, Young J, Landholder responses to incentives for wild dog control. Service Design Research Working Paper 07-11. Department of Primary Industries, Tatura, Victoria; 2011.

20. Kaine G, An application of the I3 framework to rat control in Hawke’s Bay. Landcare Research Contract Report LC3646; 2019.

21. Kaine G, Kirk N, Kannemeyer R, Stronge D, Wiercinski B. Predicting people’s motivation to engage in urban possum control. Conservation. 2021; 1: 196–215. https://doi.org/10.3390/conservation1030016.

22. Kaine G, Stronge D, An application of the I3 framework to rat control in New Plymouth. Landcare Research Report LC3734; 2020.

23. Kaine G, Kannemeyer R, Stronge D, Using 1080 to control possums and rats: An application of the I_3_ framework. Landcare Research Report LC3747; 2020.

24. New Zealand Government, History of the COVID-19 alert system; 2021. https://covid19.govt.nz/alert-system/history-of-the-covid-19-alert-system/

25. Travica B, Containment strategies for COVID-19 pandemic 2020; Available at SSRN: https://ssrn.com/abstract=3604519 or http://dx.doi.org/10.2139/ssrn.3604519

26. Ministry of Health, 2021. https://www.health.govt.nz/our-work/diseases-and-conditions/covid-19-novel-coronavirus/covid-19-response-planning/covid-19-elimination-strategy-aotearoa-new-zealand

27. Laurent G, Kapferer J.-N, Measuring consumer involvement profiles. J. Mark. Res. 1985; 22: 41–53.

28. Kaine G, A pilot application of the I_3_ framework to compliance behaviour in farming. Landcare Research Contract Report LC3513; 2019.

29. Olsen SO, Strength and conflicting valence in measurement of food attitudes and preferences. Food Quality and Preferences 1999; 10: 483–494.

30. Daoust JF, Nadeau R, Dassonneville R, Lachapelle E, Bélanger É, Savoie J, van der Linden C, How to survey citizens’ compliance with COVID-19 Public Health Measures: evidence from three survey experiments. Journal of Experimental Political Science 2020;1–8.

31. Daoust JF, Bélanger É, Dassonneville R, Lachapelle E, Nadeau R, Becher M, Brouard S, Foucault M, Hönnige C, Stegmueller D, A guilt-free strategy increases self-reported noncompliance with COVID-19 preventive measures: Experimental evidence from 12 countries. PLoS ONE. 2021; 16(4): e0249914. https://doi.org/10.1371/journal.pone.0249914

32. Aldenderfer MS, Blashfield RK, Cluster analysis. Sage, Newbury Park, California; 1984.

33. IBM Corp. IBM SPSS Statistics for Windows, Version 27.0. 2020. Armonk, NY: IBM Corporation.

34. Carmines EG, Zeller RA. Reliability and validity assessment. 1979, Newbury Park, CA: Sage.

35. Kaine G, Willingness to wear masks, self-isolate and test for Covid-19 and implications for compliance. Landcare Research Contract Report LC3867; 2020.

36. Wilson S, Pandemic leadership: Lessons from New Zealand’s approach to COVID-19. Leadership 2020; 16: 279–293. (doi:10.1177/1742715020929151)

37. Tukey J. Comparing Individual Means in the Analysis of Variance. 1949. Biometrics, 5(2), 99–114. doi:10.2307/3001913

38. Allington D, Duffy B, Wessely S, Dhavan N & Rubin J. Health-protective behaviour, social media usage, and conspiracy belief during the COVID-19 public health emergency. 2020. Psychological Medicine. https://doi.org/10.1017/S003329172000224X

39. Imhoff R, Lamberty P, A bioweapon or a hoax? The link between distinct conspiracy beliefs about the Coronavirus disease (COVID-19) outbreak and pandemic behavior. 2020. https://doi.org/10.1177/1948550620934692

40. Lieberoth A, Lin S-Y, Stöckli S, Han H, Kowal M, Gelpi R, et al., Stress and worry in the 2020 coronavirus pandemic: relationships to trust and compliance with preventive measures across 48 countries in the COVIDiSTRESS global survey, 2021, R. Soc. open sci.8200589200589, http://doi.org/10.1098/rsos.200589

41. Howard J, Huang A, Li Z, Tufekci Z, Zdimal V, van der Westhuizen H-M, et al. An evidence review of face masks against COVID-19. 2021. Proceedings of the National Academy of Sciences Jan 2021, 118 (4) e2014564118; DOI: 10.1073/pnas.2014564118

42. Gray L, MacDonald C, Tassell-Matamua N, Stanley J, Kvalsvig A, Zhang J, et al., Wearing one for the team: views and attitudes to face covering in New Zealand/Aotearoa during COVID-19 Alert Level 4 lockdown. 2020, J Prim Health Care. 2020 Sep;12(3):199–206. doi: 10.1071/HC20089. PMID: 32988441.

43. Bridgman A, Merkley E, Loewen PJ, Owen T, Ruths D, Teichmann L, et al, The causes and consequences of COVID-19 misperceptions: Understanding the role of news and social media. The Harvard Kennedy School (HKS) Misinformation Review, 2020; 1: Special Issue on COVID-19 and Misinformation.

44. Vally Z. Public perceptions, anxiety, and the perceived efficacy of health-protective behaviours to mitigate the spread of the SARS-Cov-2/COVID-19 pandemic. Public Health. 2020. doi:10.1016/j.puhe.2020.08.002

45. Taylor S, Asmundson GJG. Negative attitudes about facemasks during the COVID-19 pandemic: The dual importance of perceived ineffectiveness and psychological reactance. 2021. PLoS ONE 16(2): e0246317. https://doi.org/10.1371/journal.pone.0246317

46. Isch, C., Guevara Beltran, D., Ayers, J. D., Alcock, J., Cronk, L., Hurmuz-Sklias, H., … Aktipis, A. (2021, March 28). What predicts attitudes about mask wearing? https://doi.org/10.31234/osf.io/jvspx

47. Zhang X, Wang F, Zhu C, Wang Z. Willingness to Self-Isolate When Facing a Pandemic Risk: Model, Empirical Test, and Policy Recommendations. Int J Environ Res Public Health. 2019;17(1):197. Published 2019 Dec 27. doi:10.3390/ijerph17010197

48. Escandon-Barbosa D. Hurtado A. Gomez A. Factors Affecting Voluntary Self-Isolation Behavior to Cope with a Pandemic: Empirical Evidence from Colombia vs. Spain in Times of COVID-19. 2021. Behav. Sci., 11, 35. https://doi.org/10.3390/bs11030035

49. Lan R, Sujanto R, Lu K, He Z, Zhang CJP, Ming WK. Perceived Effectiveness, Safety, and Attitudes Toward the Use of Nucleic Tests of SARS-CoV-2 Among Clinicians and General Public in China. Front Public Health. 2020;8:599862. Published 2020 Dec 17. doi:10.3389/fpubh.2020.599862

50. Lang, R., Benham, J.L., Atabati, O. et al. Attitudes, behaviours and barriers to public health measures for COVID-19: a survey to inform public health messaging. BMC Public Health 21, 765 (2021). https://doi.org/10.1186/s12889-021-10790-0

51. West, R., Michie, S., Rubin, G.J. et al. Applying principles of behaviour change to reduce SARS-CoV-2 transmission. Nat Hum Behav 4, 451–459 (2020). https://doi.org/10.1038/s41562-020-0887-9

52. Stats NZ. Estimated resident population (ERP), subnational population by ethnic group, age, and sex, at 30 June 1996, 2001, 2006, 2013, and 2018 (2020). http://nzdotstat.stats.govt.nz/WBOS/Index.aspx?DataSetCode=TABLECODE7512#

53. Stats NZ. Highest qualification and ethnic group (grouped total responses) by age group and sex, for the census usually resident population count aged 15 years and over, 2006, 2013, and 2018 Censuses (2020). http://nzdotstat.stats.govt.nz/wbos/Index.aspx?_ga=2.69061078.636843804.1602117753761746062.1551927941#

54. Stats NZ. Household income by region, household type, and source of household income (2020). http://nzdotstat.stats.govt.nz/wbos/Index.aspx?_ga=2.69061078.636843804.1602117753-761746062.1551927941#

