## Appendix A for "Compliance with Covid-19 measures: evidence from New Zealand"

### Appendix. Summary of sample demographics

##### Table A1. Age distribution of respondents

| Age category | Proportion of respondents | Proportion of Auckland residents[1] |
|---|---|---|
| 18-29 years | 22.8 | 22.4 |
| 30-39 years | 21.8 | 20.3 |
| 40-49 years | 18.4 | 18.0 |
| 50-59 years | 13.1 | 16.7 |
| 60-69 years | 12.5 | 11.9 |
| 70 years and over | 11.4 | 10.8 |

Notes: [1] Source [52]

##### Table A2. Education distribution of respondents

| Education category | Proportion of respondents | Proportion of Auckland residents[1] |
|---|---|---|
| Some or all of secondary school | 14.2 | 23.6 |
| Certificate (1-6) | 12.4 | 35.8 |
| Diploma (5-7) | 14.3 | 9.6 |
| Graduate or post-graduate | 59.0 | 31.1 |

Notes: [1] Source [53]

##### Table A3. Ethnicity distribution of respondents

| Ethnic category | Proportion of respondents | Proportion of Auckland residents[1] |
|---|---|---|
| European | 53.3 | 47.5 |
| Māori | 4.4 | 10.3 |
| Pacific Islander | 4.7 | 14.1 |
| Other | 37.6 | 28.1 |

Notes: [1] Source [52]

##### Table A4. Income distribution of respondents

| Income category | Proportion of respondents | Proportion of Auckland residents[1] |
|---|---|---|
| Less than $20,000 | 4.3 | 8.1 |
| $20,000 to $50,000 | 21.2 | 18.7 |
| $50,000 to $70,000 | 18.6 | 11.3 |
| $70,000 to 4100,000 | 22.0 | 14.7 |
| More than $100,000 | 33.8 | 47.0 |

Notes: [1] Source [54]
