## Supplementary material for "Compliance with Covid-19 measures: evidence from New Zealand": S1 Questionnaire

### Covid19 Survey Questionnaire

SAMPLE: 1000 households in Auckland:

- 333 respondents answer Q5-Q9 and Q10-Q15 (Group A)
- 333 respondents answer Q10-Q15 and Q16-Q21 (Group B)
- 333 respondents answer Q5-Q9 and Q16-Q21 (Group C)
- All respondents answer Q1-Q4 and Part B.

**Q1: Beliefs about Covid-19**

We are interested in your opinions about Covid-19. How strongly do you agree or disagree with the following statements?

| Item | Strongly agree | Agree | Unsure/ neutral | Disagree | Strongly disagree |
|---|---|---|---|---|---|
| You cannot catch Covid-19 from people with the virus who do not have symptoms | ☐ | ☐ | ☐ | ☐ | ☐ |
| Covid-19 is only a danger to the elderly and people who already have health problem | ☐ | ☐ | ☐ | ☐ | ☐ |
| Infected people spread Covid-19 by coughing and sneezing | ☐ | ☐ | ☐ | ☐ | ☐ |
| Children cannot catch Covid-19 | ☐ | ☐ | ☐ | ☐ | ☐ |
| Once you have had Covid-19 you are immune to re-infection | ☐ | ☐ | ☐ | ☐ | ☐ |
| I think Covid-19 is a hoax | ☐ | ☐ | ☐ | ☐ | ☐ |
| Fears about Covid-19 are exaggerated | ☐ | ☐ | ☐ | ☐ | ☐ |
| Covid-19 most likely comes from bats | ☐ | ☐ | ☐ | ☐ | ☐ |
| Covid-19 is a man-made virus | ☐ | ☐ | ☐ | ☐ | ☐ |
| Children are perfectly safe from Covid-19 | ☐ | ☐ | ☐ | ☐ | ☐ |
| You can catch Covid-19 by touching anything handled by an infected person | ☐ | ☐ | ☐ | ☐ | ☐ |
| Covid-19 is no worse than the seasonal flu | ☐ | ☐ | ☐ | ☐ | ☐ |

**Q2: Beliefs about eliminating Covid-19**

We are interested in your opinions about eliminating Covid-19. How strongly do you agree or disagree with the following statements?

| Item | Strongly agree | Agree | Unsure/ neutral | Disagree | Strongly disagree |
|---|---|---|---|---|---|
| We need to eliminate Covid-19 from New Zealand to save lives | ☐ | ☐ | ☐ | ☐ | ☐ |
| We should just live with it until we have a vaccine | ☐ | ☐ | ☐ | ☐ | ☐ |
| It would be better to let it spread and build herd immunity | ☐ | ☐ | ☐ | ☐ | ☐ |
| There is no point trying to eliminate Covid-19 because it is a virus and will keep changing | ☐ | ☐ | ☐ | ☐ | ☐ |
| Covid-19 is everywhere in the world so there is no way we can keep it out | ☐ | ☐ | ☐ | ☐ | ☐ |

**Q3: Taking responsibility and action**

We are interested in how strongly you feel about the need for action to be taken to eliminate Covid-19 from New Zealand. How strongly do you agree or disagree with the following statements?

| Item | Strongly agree | Agree | Unsure/ neutral | Disagree | Strongly disagree |
|---|---|---|---|---|---|
| Eliminating Covid-19 from New Zealand is the right thing to do | ☐ | ☐ | ☐ | ☐ | ☐ |
| I feel some responsibility for eliminating Covid-19 from New Zealand | ☐ | ☐ | ☐ | ☐ | ☐ |
| I am prepared to change my normal behaviour to eliminate Covid-19 from New Zealand | ☐ | ☐ | ☐ | ☐ | ☐ |
| It is important to work together to eliminate Covid-19 from New Zealand | ☐ | ☐ | ☐ | ☐ | ☐ |
| Nearly everyone I know thinks eliminating Covid-19 from New Zealand is the right thing to do | ☐ | ☐ | ☐ | ☐ | ☐ |
| Most people I know feel some responsibility for eliminating Covid-19 from New Zealand | ☐ | ☐ | ☐ | ☐ | ☐ |
| I think nearly everyone is prepared to change their normal behaviour to eliminate Covid-19 from New Zealand | ☐ | ☐ | ☐ | ☐ | ☐ |
| I am prepared to make sacrifices to eliminate Covid-19 from New Zealand | ☐ | ☐ | ☐ | ☐ | ☐ |
| Most people are prepared to make sacrifices to eliminate Covid-19 from New Zealand | ☐ | ☐ | ☐ | ☐ | ☐ |
| Most people know we must work together to eliminate Covid-19 from New Zealand | ☐ | ☐ | ☐ | ☐ | ☐ |

**Q4: Involvement with eliminating Covid-19 from New Zealand**

We are interested in your opinions about eliminating Covid-19 from New Zealand. How strongly do you agree or disagree with the following statements?

| Item | Strongly agree | Agree | Unsure/ neutral | Disagree | Strongly disagree |
|---|---|---|---|---|---|
| I think helping to eliminate Covid-19 from New Zealand is rewarding | ☐ | ☐ | ☐ | ☐ | ☐ |
| The consequences are serious if we don't eliminate Covid-19 from New Zealand | ☐ | ☐ | ☐ | ☐ | ☐ |
| Eliminating Covid-19 from New Zealand is something I am passionate about | ☐ | ☐ | ☐ | ☐ | ☐ |
| It would be a big deal if government made mistakes while we try to eliminate Covid-19 from New Zealand | ☐ | ☐ | ☐ | ☐ | ☐ |
| My position on eliminating Covid-19 from New Zealand tells others something about me | ☐ | ☐ | ☐ | ☐ | ☐ |
| Eliminating Covid-19 from New Zealand is important to me | ☐ | ☐ | ☐ | ☐ | ☐ |
| Making decisions about how to eliminate Covid-19 from New Zealand is complicated | ☐ | ☐ | ☐ | ☐ | ☐ |
| What others think about eliminating Covid-19 from New Zealand tells me something about them | ☐ | ☐ | ☐ | ☐ | ☐ |
| I care a lot about eliminating Covid-19 from New Zealand | ☐ | ☐ | ☐ | ☐ | ☐ |
| Making decisions about how to eliminate Covid-19 from New Zealand is difficult | ☐ | ☐ | ☐ | ☐ | ☐ |

**Q5: Involvement with wearing face masks to help eliminate Covid-19?**

The government may require you to wear a face mask in public as one measure to eliminate Covid-19 from New Zealand. How strongly do you agree or disagree with the following statements about wearing face masks?

| Item | Strongly agree | Agree | Unsure/ neutral | Disagree | Strongly disagree |
|---|---|---|---|---|---|
| I think it's rewarding to wear a face mask to help eliminate Covid-19 | ☐ | ☐ | ☐ | ☐ | ☐ |
| The consequences are serious if I made mistakes with wearing a face mask to help eliminate Covid-19 | ☐ | ☐ | ☐ | ☐ | ☐ |
| Wearing a face mask to help eliminate Covid-19 is something I am passionate about | ☐ | ☐ | ☐ | ☐ | ☐ |
| It would be a big deal if I made a mistake with wearing a face mask to help eliminate Covid-19 | ☐ | ☐ | ☐ | ☐ | ☐ |
| My position about wearing a face mask to help eliminate Covid-19 tells others something about me | ☐ | ☐ | ☐ | ☐ | ☐ |
| Wearing a face mask to help eliminate Covid-19 is important to me | ☐ | ☐ | ☐ | ☐ | ☐ |
| Making decisions about wearing a face mask to help eliminate Covid-19 is complicated | ☐ | ☐ | ☐ | ☐ | ☐ |
| What others think about wearing a face mask to help eliminate Covid-19 tells me something about them | ☐ | ☐ | ☐ | ☐ | ☐ |
| I care a lot about wearing a face mask to help eliminate Covid-19 | ☐ | ☐ | ☐ | ☐ | ☐ |
| Making decisions about wearing a face mask to help eliminate Covid-19 is difficult | ☐ | ☐ | ☐ | ☐ | ☐ |

**Q6: Attitude towards wearing a face mask to stop the spread Covid-19**

How strongly do you agree or disagree with the following statements about wearing face masks to stop the spread of Covid-19?

| Item | Strongly agree | Agree | Unsure/ neutral | Disagree | Strongly disagree |
|---|---|---|---|---|---|
| I think face masks should be worn to help stop the spread of Covid-19 | ☐ | ☐ | ☐ | ☐ | ☐ |
| I think wearing face masks to stop the spread of Covid-19 is the right thing to do | ☐ | ☐ | ☐ | ☐ | ☐ |
| I believe it is wrong to wear face masks to stop the spread of Covid-19 | ☐ | ☐ | ☐ | ☐ | ☐ |
| I think it would be good to wear face masks to stop the spread of Covid-19 | ☐ | ☐ | ☐ | ☐ | ☐ |

**Q7:** Which one of the following statements best describes you?

Please choose one

| Item | Describes me |
|---|---|
| I really think wearing face masks is the right thing to do | ☐ |
| It doesn't really matter to me whether or not I wear a face mask | ☐ |
| I am not really sure if wearing face masks is the best way to go | ☐ |
| I haven't put much thought into wearing face masks | ☐ |
| I strongly believe that wearing face masks is a bad thing to do | ☐ |

**Q8: Perceived advantages and disadvantages of wearing face masks to help stop the spread of Covid-19**

How strongly do you agree or disagree with the following statements about wearing face masks to help stop the spread of Covid-19?

| Item | Strongly agree | Agree | Unsure/ neutral | Disagree | Strongly disagree |
|---|---|---|---|---|---|
| Face masks are effective in preventing the spread of Covid-19 | ☐ | ☐ | ☐ | ☐ | ☐ |
| Wearing face masks to stop the spread of Covid-19 is just not practical | ☐ | ☐ | ☐ | ☐ | ☐ |
| Face masks are not much help in stopping the spread of Covid-19 because people do not wear them properly | ☐ | ☐ | ☐ | ☐ | ☐ |
| Face masks on their own are not much help in preventing the spread of Covid-19 | ☐ | ☐ | ☐ | ☐ | ☐ |
| You should only have to wear a face mask if you feel unwell | ☐ | ☐ | ☐ | ☐ | ☐ |
| You should only have to wear a face mask if you are old or have a health problem | ☐ | ☐ | ☐ | ☐ | ☐ |
| Face masks are not much help unless you wear gloves as well | ☐ | ☐ | ☐ | ☐ | ☐ |
| Home-made face masks are a waste of time and effort | ☐ | ☐ | ☐ | ☐ | ☐ |
| Face masks are just too uncomfortable | ☐ | ☐ | ☐ | ☐ | ☐ |
| The kind of face masks we can buy are not worth bothering with | ☐ | ☐ | ☐ | ☐ | ☐ |
| Wearing face mask sets a good example to others | ☐ | ☐ | ☐ | ☐ | ☐ |
| People who wear face masks are over-reacting | ☐ | ☐ | ☐ | ☐ | ☐ |
| Wearing face masks should be compulsory | ☐ | ☐ | ☐ | ☐ | ☐ |
| Face masks are too difficult and inconvenient if you wear glasses | ☐ | ☐ | ☐ | ☐ | ☐ |

**Q9a: Did you wear a face mask whenever you went out in public last week?**

Always Often Sometimes Rarely Never NA

**Q9b: Did you wear a face mask if you had to go out to work last week?**

Always Often Sometimes Rarely Never NA

**Q10: Involvement with self-isolating to help eliminate Covid-19?**

Staying at home if you feel unwell is one strategy the government is using to help eliminate Covid-19 from New Zealand. How strongly do you agree or disagree with the following statements about staying at home if you feel unwell?

| Item | Strongly agree | Agree | Unsure/ neutral | Disagree | Strongly disagree |
|---|---|---|---|---|---|
| I think staying at home if you feel unwell to help eliminate Covid-19 would be rewarding | ☐ | ☐ | ☐ | ☐ | ☐ |
| The consequences would be serious if I made a mistake about staying at home if I felt unwell | ☐ | ☐ | ☐ | ☐ | ☐ |
| I am passionate about staying at home if I feel unwell | ☐ | ☐ | ☐ | ☐ | ☐ |
| Making mistakes about staying at home if you are feeling unwell are a big deal | ☐ | ☐ | ☐ | ☐ | ☐ |
| My position about staying at home if I feel unwell tells others something about me | ☐ | ☐ | ☐ | ☐ | ☐ |
| Staying at home if I feel unwell is important to me | ☐ | ☐ | ☐ | ☐ | ☐ |
| Making decisions about staying at home if I feel unwell is complicated | ☐ | ☐ | ☐ | ☐ | ☐ |
| What others think about staying at home if they feel unwell tells me something about them | ☐ | ☐ | ☐ | ☐ | ☐ |
| I care a lot about the need to stay home if I feel unwell | ☐ | ☐ | ☐ | ☐ | ☐ |
| Making decisions about staying at home if I feel unwell is difficult | ☐ | ☐ | ☐ | ☐ | ☐ |

**Q11: Attitude towards staying at home if you were unwell**

How strongly do you agree or disagree with the following statements about staying at home if you feel unwell?

| Item | Strongly agree | Agree | Unsure/ neutral | Disagree | Strongly disagree |
|---|---|---|---|---|---|
| I think people should stay at home if they feel unwell | ☐ | ☐ | ☐ | ☐ | ☐ |
| I think staying at home if you feel unwell is the right thing to do | ☐ | ☐ | ☐ | ☐ | ☐ |
| I believe it is wrong to stay at home if you feel unwell | ☐ | ☐ | ☐ | ☐ | ☐ |
| I think it is a good thing if people who feel unwell stay at home | ☐ | ☐ | ☐ | ☐ | ☐ |

**Q12:** Which of the following statements best describes you?

Please choose one

| Item | Describes me |
|---|---|
| I really think staying at home if you feel unwell is the right thing to do | ☐ |
| It doesn't really matter to me whether or not people stay at home if they feel unwell | ☐ |
| I am not really sure that staying at home if you feel unwell is the best way to go | ☐ |
| I haven't put much thought into staying at home if you feel unwell | ☐ |
| I strongly believe that staying at home if you feel unwell is a bad thing to do | ☐ |

#### Q13: Perceived advantages and disadvantages of staying at home if you feel unwell

How strongly do you agree or disagree with the following statements about staying at home if you feel unwell?

| Item | Strongly agree | Agree | Unsure/ neutral | Disagree | Strongly disagree |
|---|---|---|---|---|---|
| Staying at home if you feel unwell is effective in preventing the spread of Covid-19 | ☐ | ☐ | ☐ | ☐ | ☐ |
| Staying at home if you feel unwell is just not practical | ☐ | ☐ | ☐ | ☐ | ☐ |
| I think staying at home if you were unwell would be depressing | ☐ | ☐ | ☐ | ☐ | ☐ |
| I couldn't afford the time off work to stay home if I was unwell | ☐ | ☐ | ☐ | ☐ | ☐ |
| You should only stay at home if you have all the right symptoms | ☐ | ☐ | ☐ | ☐ | ☐ |
| You should only have to stay at home if you are old or already have a health problem | ☐ | ☐ | ☐ | ☐ | ☐ |
| Staying home if you are unwell is not much help if you don't get tested | ☐ | ☐ | ☐ | ☐ | ☐ |
| Staying at home if you feel unwell is a waste of time and effort | ☐ | ☐ | ☐ | ☐ | ☐ |

#### Q14: Thinking about the next few days, would you stay home if you were unwell or have any of the following symptoms: a dry cough, fever, loss of sense of smell, loss of sense of taste, shortness of breath or difficulty breathing?

| Definitely | Probably | Maybe | Probably not | Definitely not |
|---|---|---|---|---|
| ☐ | ☐ | ☐ | ☐ | ☐ |

#### Q15: If you were advised to do so by a healthcare professional or public health authority would you self-isolate for 14 days?

| Definitely | Probably | Maybe | Probably not | Definitely not |
|---|---|---|---|---|
| ☐ | ☐ | ☐ | ☐ | ☐ |

**Q16: Involvement with testing to help eliminate Covid-19?**

Testing for Covid-19 is one strategy the government is using to help eliminate the virus from New Zealand. How strongly do you agree or disagree with the following statements about testing?

| Item | Strongly agree | Agree | Unsure/ neutral | Disagree | Strongly disagree |
|---|---|---|---|---|---|
| I think getting tested to help eliminate Covid-19 is rewarding | ☐ | ☐ | ☐ | ☐ | ☐ |
| The consequences are serious if I make a mistake about getting tested for Covid-19 | ☐ | ☐ | ☐ | ☐ | ☐ |
| Getting tested for Covid-19 is something I am passionate about | ☐ | ☐ | ☐ | ☐ | ☐ |
| It would be a big deal if I made a mistake with getting tested for Covid-19 | ☐ | ☐ | ☐ | ☐ | ☐ |
| My position about getting tested for Covid-19 tells others something about me | ☐ | ☐ | ☐ | ☐ | ☐ |
| Getting tested for Covid-19 is important to me | ☐ | ☐ | ☐ | ☐ | ☐ |
| Making decisions about getting tested for Covid-19 is complicated | ☐ | ☐ | ☐ | ☐ | ☐ |
| What others think about getting tested for Covid-19 tells me something about them | ☐ | ☐ | ☐ | ☐ | ☐ |
| I care a lot about getting tested for Covid-19 | ☐ | ☐ | ☐ | ☐ | ☐ |
| Making decisions about getting tested for Covid-19 is difficult | ☐ | ☐ | ☐ | ☐ | ☐ |

**Q17: Attitude towards getting tested for Covid-19**

How strongly do you agree or disagree with the following statements about getting testing for Covid-19?

| Item | Strongly agree | Agree | Unsure/ neutral | Disagree | Strongly disagree |
|---|---|---|---|---|---|
| I think people should get tested for Covid-19 | ☐ | ☐ | ☐ | ☐ | ☐ |
| I think getting tested for Covid-19 is the right thing to do | ☐ | ☐ | ☐ | ☐ | ☐ |
| I believe it is wrong to test people for Covid-19 | ☐ | ☐ | ☐ | ☐ | ☐ |
| I think it is good to test people for Covid-19 | ☐ | ☐ | ☐ | ☐ | ☐ |

**Q18:** Which of the following statements best describes you?

Please choose one

| Item | Describes me |
|---|---|
| I really think testing people for Covid-19 is the right thing to do | ☐ |
| It doesn't really matter to me whether or not people are tested for Covid-19 | ☐ |
| I am not really sure if testing people for Covid-19 is the best way to go | ☐ |
| I haven't put much thought into testing people for Covid-19 | ☐ |
| I strongly believe that testing people for Covid-19 is a bad thing to do | ☐ |

**Q19: Perceived advantages and disadvantages of testing people for Covid-19**

How strongly do you agree or disagree with the following statements about testing people for Covid-19?

| Item | Strongly agree | Agree | Unsure/ neutral | Disagree | Strongly disagree |
|---|---|---|---|---|---|
| Testing people is effective in preventing the spread of Covid-19 | ☐ | ☐ | ☐ | ☐ | ☐ |
| Testing people to stop the spread of Covid-19 is just not is not practical | ☐ | ☐ | ☐ | ☐ | ☐ |
| The tests for Covid-19 are too unreliable | ☐ | ☐ | ☐ | ☐ | ☐ |
| I couldn't afford the time off work if I tested positive | ☐ | ☐ | ☐ | ☐ | ☐ |
| You should only get tested if you have the right symptoms | ☐ | ☐ | ☐ | ☐ | ☐ |
| You should only get tested if you are old or have a health problem | ☐ | ☐ | ☐ | ☐ | ☐ |
| Testing is not much help if you only test sick people | ☐ | ☐ | ☐ | ☐ | ☐ |
| Getting tested is a waste of time and effort | ☐ | ☐ | ☐ | ☐ | ☐ |
| Testing is painful and uncomfortable | ☐ | ☐ | ☐ | ☐ | ☐ |
| Testing takes so long it's not worth bothering with | ☐ | ☐ | ☐ | ☐ | ☐ |

**Q20: Have you been tested for Covid-19?** **Yes**

**No**

If answer to Q20 is 'Yes' then ask Q21, otherwise go to B1

**Q21: Did you feel unwell at the time you were tested?** **Yes**

**No**

**Part B: Demographics**

**B1: What age bracket do you fit into?**

( ) 18-29 years

( ) 30-39 years

( ) 40-49 years

( ) 50-59 years

( ) 60-69 years

( ) 70 years and over

( ) Prefer not to say

**B2: Which of the following do you identify as?**

( ) Male

( ) Female

( ) Gender diverse

( ) Prefer not to say

**B3: What is your highest level of formal education?**

( ) Some or all of secondary school

( ) Certificate (1-6)

( ) Diploma (5-7)

( ) Bachelor degree

( ) Post-graduate diploma/certificate

( ) Post-graduate degree

( ) Prefer not to say

**B4: What is your ethnicity?**

( ) Māori

( ) European New Zealander

( ) Pacific Islander

( ) Asian

( ) Other

**B5: What household income bracket do you fit into?**

( ) Less than $20,000 etc based on census scales

( ) $20,000 to $50,000

( )$50,000 to $70,000

( ) $70,000 to $100,000

( ) more than $100,000

( ) Prefer not to say
